# RadKev: An Open-Weight Decision Model for Radiology

**DOI:** 10.64898/2026.10.07.26364991

**Authors:** Udbhav Ram, Ran Zhang

## Abstract

**Background:** Many radiology decisions, such as whether a report describes a finding or how urgently a study should be read, are choices among a fixed set of answers. Decision models return a probability for each permitted answer in one step, without generating text, and an open-weight model can run within a hospital so that patient data stay local. No decision model has yet been specialized for radiology and compared with large language models (LLMs).

**Methods:** We fine-tuned the general-purpose decision models Kev-27B (Qwen3.8-27B backbone) and Kev-9B on 36,109 records from public radiology datasets to obtain RadKev-27B and RadKev-9B, and compared them with the Kev models, the hosted decision models Jev and OpenAI Decisions, and the LLM Qwen3.8-27B, on 14,142 held-out questions in fifteen tasks (report reading, case diagnosis and classification, radiological knowledge). The prespecified primary outcome was task-averaged accuracy relative to Kev-27B.

**Results:** RadKev-27B exceeded Kev-27B by 4.9 percentage points (95% CI 3.8 to 6.2), with the largest gain in report error detection (22.6 points); RadKev-9B exceeded Kev-9B by 6.9 points (5.5 to 8.4) and did not differ detectably from Kev-27B, a model three times its size. RadKev-27B was more accurate than the hosted decision models Jev (3.0 points, 1.7 to 4.4) and OpenAI Decisions (4.8, 3.2 to 6.4), than Qwen3.8-27B without reasoning (4.7, 3.4 to 6.0) and with reasoning (2.8 points, 1.0 to 4.6) on 1,675 questions, at 132-fold lower latency (128 ms vs 16.9 s per question). At an error rate of 5%, RadKev-27B could answer 90.5% of questions and Kev-27B 76.4%.

**Conclusions:** RadKev-27B was more accurate than two hosted decision models and an LLM of the same size, with or without reasoning, at a fraction of the latency of reasoning, and can be deployed within an institution for categorical radiology decisions after validation on local data.

## 1 Introduction

### 1.1 Categorical decisions in radiology

Many routine radiology workflows involve categorical decision making: whether a report describes a particular finding, which differential diagnosis within a specific subset of options is most likely, to which subspecialty a study should be assigned, and how urgently a result requires triage and follow-up. Increasing automation in such decisions requires a model that selects consistently and confidently among predefined answers and assigns a probability to its selection, so that highly confident decisions can be automated and uncertain ones can be flagged for human review [1]. Large language models (LLMs) have been evaluated for many such decisions, including the extraction of findings from free-text reports [2, 3], the detection of errors in reports [4], the selection of imaging according to the American College of Radiology (ACR) Appropriateness Criteria [5, 6], diagnosis from the clinical history and imaging findings [7, 8] and radiology board-style questions [9]. Three properties of LLMs limit their use for such decisions. First, an LLM returns generated text, which must be parsed and mapped onto the admissible answers and does not directly provide a probability for each answer. Second, text is generated sequentially, one token at a time, so that latency and compute increase with the length of the output, particularly when the model reasons before answering. Third, the most capable LLMs are often externally hosted and are therefore unsuitable for patient reports under institutional privacy constraints [3].

### 1.2 Decision models

Decision models, introduced by TypeSafe as so-called System One models, are designed for this setting [10]. A decision model receives an input text, the state, together with typed questions (binary, multiple-choice or ordinal) whose admissible answers are specified in the request body, and returns a probability distribution over the admissible answers to each question. All questions are answered in a single query, in parallel, rather than by autoregressive generation [10, 11], so every output is a valid answer by construction. Jev, released by TypeSafe in September 2026, is available only through an externally hosted interface, so that submitted data must leave the user’s institution, and its architecture remains closed source [11, 12]. The OpenAI Decisions API, released in public beta in October 2026, likewise provides a general-purpose decision model through an externally hosted interface [13].

Kev is an open-weight family of decision models, with 0.8 to 27 billion parameters, that implements the same TypeSafe-like interface on open-source Qwen backbones [14], with an architecture inferred from Jev’s external behavior [12, 15]. Because its weights and training code are released under the Apache-2.0 license, Kev can be deployed within an institutional network and adapted to domain-specific data. These properties correspond closely to the requirements of radiology: the decisions are categorical and frequent, the several decisions required for one report can be answered in a single request, calibrated probabilities permit selective automation, and patient data need not leave the institution.

### 1.3 Domain specialization

In a benchmark evaluation, a general-purpose decision model, Jev, achieved accuracy comparable to that of the frontier LLM GPT-6 Sol with reasoning on questions derived from research abstracts, but lower accuracy on examination questions and substantially lower accuracy on diagnostic case challenges (59.8% vs 82.4% on DiagnosisArena), although its probabilities were better calibrated on examination questions [16]. In radiology, Jev has been used to evaluate the factual consistency of generated reports [17]. For LLMs, the value of domain specialization is contested: domain-adaptive pretraining improves downstream performance [18], whereas a generalist model with optimized prompting has exceeded medically fine-tuned models [19]. For decision models, domain adaptation has been reported for Chinese-language medical, legal and financial tasks [20]. To our knowledge, no decision model has been specialized for radiology, and specialization has not been compared with the principal alternatives for direct deployment in radiology practice: larger general-purpose decision models and LLMs of comparable size.

### 1.4 Present study

In this study, we fine-tuned two general-purpose decision models, Kev-27B and Kev-9B, on 36,109 records from public radiology sources to obtain RadKev-27B and RadKev-9B, and evaluated them on a benchmark of 14,142 held-out questions on decisions of the kinds for which LLMs have been evaluated in radiology, under an analysis plan specified before any result of these models was examined. We tested the hypothesis that a decision model specialized for radiology makes these decisions at least as accurately as general-purpose decision models and as an LLM of comparable size, at a fraction of the inference latency and compute of an LLM that generates its answers, because it answers every question about a record in a single forward pass without generating text. Specialization was compared with model scale, represented by the general-purpose decision models Kev-9B and Kev-27B, and with an LLM of the same size, Qwen3.8-27B [14], evaluated zero-shot with reasoning disabled on every question it could answer and enabled on a sample of them. The specialized models were also compared with two externally hosted general-purpose decision models, Jev and the OpenAI Decisions API, and evaluated on an external test set of radiology reports from another institution [21]. Because Kev-27B is built on Qwen3.8-27B, the design compares identical backbone weights used as an LLM, as a general-purpose decision model and as a radiology-specialized decision model. The weights of RadKev-27B and RadKev-9B, the training and evaluation code and the benchmark, where source licenses permit, are publicly released.

## 2 Methods

### 2.1 Data

The data used for this study are intended to cover three decision-making modalities in radiology for which openly available data exist: reading a report, diagnosing or classifying a case and applying radiological knowledge (Table 1). Each record pairs a state, i.e. the input text that the model ingests, with one or more typed questions whose admissible answers are given with the question: yes or no, a choice among named options, or an ordinal score. Ground truth generated or verified by human annotators was preferred wherever it was available; the datasets whose ground truth is not human-generated, namely ReXErr (known by construction), CT-RATE (a report classifier) and the questions on orders, triage and follow-up (agreement of two LLMs), are identified below. Records were divided into training, development and test splits by patient or case, with official test sets retained where available, and one instruction wording per question kind was withheld from training. Development records were never used to update the model weights; they served to monitor training and to fit the temperature of each model (Section 2.3). IU/Open-i, MMLU, PubMedQA and MedXpertQA therefore contributed development and test records but no training records: the license of IU/Open-i does not permit derivative works, and MMLU, the expert-annotated subset of PubMedQA and MedXpertQA are distributed as evaluation collections, which also places sources not seen in training in the benchmark. Supplementary Note S1 shows example records of every dataset, and Supplementary Note S2 gives the construction rules.

**Table 1.** Data used in this study. Train and Dev, records used for training and development; Test, questions held out of training; a dash indicates no records. The radiology benchmark comprises 14,142 of the test questions. The others are the classifier-labeled structured label extraction questions, the LLM-labeled questions on orders, triage and follow-up, and the external test: radiologist-annotated CT and MRI reports from another institution, analyzed separately from the benchmark to assess generalization. Section 2.1 describes each dataset. Case text © European Society of Radiology, reproduced under CC BY-NC-SA 4.0; this figure is distributed under the same license.

| Dataset | Train | Dev | Test |
| --- | --- | --- | --- |
| Structured label extraction | 8,000 | 2,797 | 16,168 |
| Report error detection | 6,000 | 500 | 2,708 |
| Case diagnosis and classification | 2,912 | 329 | 610 |
| Imaging appropriateness | 264 | 29 | 227 |
| Radiology knowledge | 7,490 | 1,644 | 1,379 |
| Orders, triage and follow-up | 11,443 | 877 | 1,025 |
| External test | – | – | 4,037 |
| Total | 36,109 | 6,176 | 26,154 |

#### Structured label extraction

These questions ask which findings a radiology report describes. IU/Open-i [22] comprises chest radiograph reports whose findings were indexed by human coders with Medical Subject Headings; the indexed terms were mapped to 14 findings, and each report was posed yes/no questions on whether a finding is present and whether the study is normal and a four-option question on which finding the report describes, with the human indexing as the ground truth. IU/Open-i was used only for development and testing. CT-RATE [23] comprises chest CT reports posed in the same way for 18 abnormalities, whose labels were produced by a RadBERT report classifier fine-tuned on manually annotated reports; because its ground truth is not human-generated, CT-RATE was used for training, and its held-out test questions are reported as agreement with the classifier rather than as part of the benchmark. RadGraph-XL [21], whose reports were annotated by radiologists, served as the external test set (Section 2.6).

#### Report error detection

ReXErr [24] pairs MIMIC-CXR reports [25] with versions into which GPT-4o inserted clinically meaningful errors. Each record shows either the original or the error-injected version of a report, and the question asks whether the report contains an error; the ground truth, whether an error was inserted, is therefore known by construction rather than assigned by a reader. The official splits were used, with 6,000 training and 500 development reports sampled and all 2,708 test reports retained (1,376 with an injected error).

#### Case diagnosis and classification

Eurorad [26, 27] comprises teaching cases written by radiologists and published by the European Society of Radiology. The state contains the patient’s age, sex, clinical history and imaging findings, from which sentences revealing the diagnosis were removed, and the model selects the author’s final diagnosis from the case’s own differential diagnoses or the subspecialty section, among 11, under which the case was published; both ground truths were assigned by the case authors and editors. RSNA-RadioQA [8] comprises case descriptions from the Case Collection of the Radiological Society of North America (RSNA), posed as a choice among four diagnoses. Of its 80 published questions, the 79 whose text appears in the supplement of the original study were used, with the four answer options of the RaR study [28], whose incorrect options were generated by LLMs and reviewed by a board-certified radiologist, and with the published reference answer as the ground truth. RSNA-RadioQA was used only for testing.

#### Imaging appropriateness

RadCases [6] comprises single-sentence patient summaries labeled with the ACR Appropriateness Criteria panel and topic that govern their imaging work-up; the labels were assigned by two senior medical students under the supervision of an attending radiologist, with disagreements adjudicated by radiologists. Each summary was posed as a choice among 12 panel options (the panels and an option stating that no topic applies) and among 225 topics. Only the openly available subsets were used, summaries written by GPT-3.5 and the first sentences of Medbullets USMLE Step 2 and 3 questions [29]: 468 cases were matched to their labels, 37 whose text occurred in another split of this study were excluded, and the remainder were split by case into 264 training, 29 development and 138 test cases.

#### Radiological knowledge

Examination questions, whose answers were set by the authors of the examinations, measure the radiological knowledge on which the clinical decisions rely, although they are not clinical tasks themselves. To focus training and evaluation on radiology, the MedMCQA [30] radiology subject was used in full, and from the other MedMCQA subjects, MedQA [31], the medical subjects of MMLU [32], the expert-annotated subset of PubMedQA [33] and MedXpertQA [34] only the questions whose stem, state or options contain a term from a fixed list of imaging modalities and radiological terms were used (Supplementary Note S2). The same selection was applied to all splits; it retained 1,379 of 8,995 test questions and, for training, 5,070 of 35,634 MedMCQA and 2,420 of 9,175 MedQA records. MedMCQA and MedQA contributed training, development and test questions, whereas MMLU, PubMedQA and MedXpertQA were used only for development and testing.

#### Orders, triage and follow-up

Decisions about imaging orders, triage and follow-up have no public ground truth. Questions of this kind were therefore generated from public reports and clinical indications and labeled by the agreement of two LLMs (Section 2.2); they were used only to augment the training data, and their held-out test questions were not evaluated.

The benchmark thus measures performance on clinically relevant radiology decisions, together with the underlying radiological knowledge they require. It comprises the 14,142 held-out test questions of the datasets above other than CT-RATE, RadGraph-XL and the LLM-labeled questions: 11,434 whose groundtruth labels were generated or verified by human annotators (human-labeled questions) and the 2,708 ReXErr questions, whose ground truth is known by construction.

### 2.2 LLM-labeled training data

Decisions about imaging orders, triage and follow-up are routine in radiology, but no large-scale public datasets provide reliable human-generated ground-truth labels for them, and labeling them manually at the required scale was not feasible within this study. Questions on these decisions were therefore labeled by LLMs and used as a form of training-data augmentation: they extend the training data to decisions that have no public ground-truth labels, transferring the judgments of two teacher models to the decision model as in knowledge distillation [35], and they were not used for testing. Candidate questions on these decisions, of ten kinds (Supplementary Table S1), were generated from reports in CheXpert Plus [36], ReXGradient160K [37] and CT-RATE and from clinical indications in these sources and in Eurorad. Two LLMs acted as teachers (Figure 1): MedGemma-27B-text [38], a medical model, and Qwen3.8-27B [14], a general-purpose model. Each read the state and the question with lettered options, with reasoning disabled, and its answer distribution was the softmax of its next-token logits over the option letters. In the labels used for training, the letter probabilities of MedGemma-27B-text were read at the start of the reasoning segment that it opens by default rather than after it; when the labels were regenerated with the segment closed, 93.1% of the retained training questions were retained again and none received a different label (Supplementary Note S2). A question was retained only if both teachers ranked the same answer option first; the agreed-upon answer became the label, and the mean of the two distributions a soft training target [35]. Of 33,000 candidates, 22,557 (68.4%) were retained, 19,945 of them in the training split. Machine-derived labels are widely used in radiology datasets [23, 39], LLM annotation has performed comparably to crowd-worker annotation on text-classification tasks [40], and agreement between independent labelers is an established filter for label errors [41]. Because LLM labels still require validation against human judgment [42], and because labels produced by Qwen3.8-27B, which is also a comparator, would favor that model, the teacher-labeled questions were used only for training and are not part of the benchmark.

**Figure 1.**
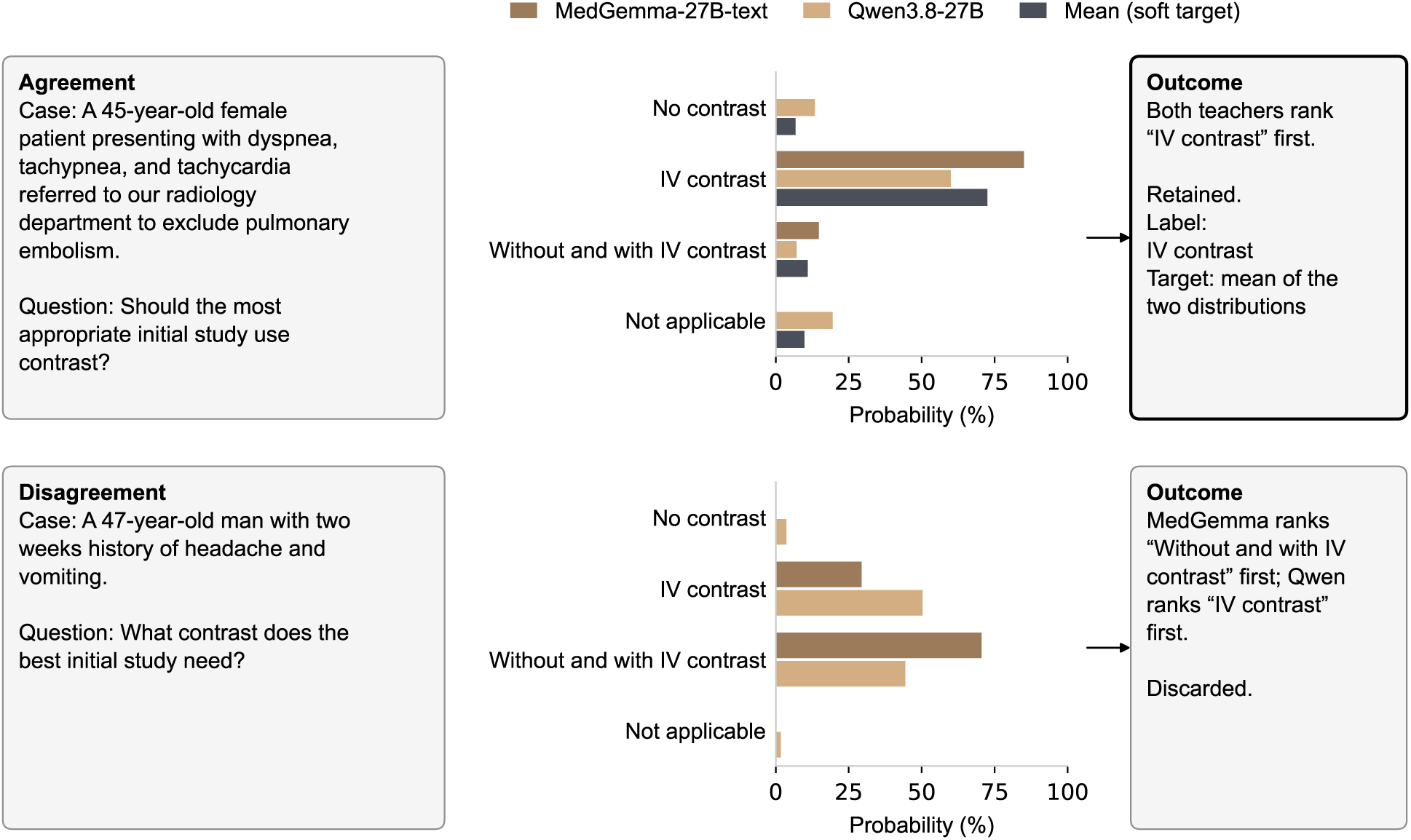
Two-teacher labeling, illustrated with two order questions generated from Eurorad case presentations (cases 17094 and 18086). Each teacher’s answer distribution is the softmax of its next-token logits over the option letters. When both teachers rank the same option first (top), the question is retained, with that option as its label and the mean of the two distributions as its soft training target; otherwise (bottom), it is discarded and receives no soft target.

### 2.3 Model and training

Kev [15] adds a low-rank adapter (LoRA) [43] of rank 16 to every linear projection of a frozen language-model backbone, together with a pointer head [44] (Figure 2): the state is encoded once, each question attends to the state but not to the other questions, and the resultant answer distribution is a softmax over scores computed between the question and each of its options. RadKev-27B and RadKev-9B were obtained by continued training of the released Kev-27B and Kev-9B (revisions 01b8199 and 2629c06), whose backbones are Qwen3.8-27B and Qwen3.5-9B-Base, so that the specialized models retain Kev’s interface and differ from it only in their weights. The training split of all sources in Table 1 (36,109 records) was interspersed with 1,000 records from Kev’s own training data to limit the loss of general decision skill (of these 37,109 records, 155 exceeded the training context of Kev’s trainer, a state of 1,536 tokens, a question branch of 2,176 tokens or a packed record of 3,200 tokens, and were excluded at loading, leaving 36,954), and Kev’s prescribed augmentations were applied: options were reshuffled in each pass, and questions with a single correct answer were randomly modified with “none of the above” options or irrelevant distractors, whereas teacher-labeled questions were only reshuffled to preserve their soft targets. The loss was the cross-entropy with respect to the labeled answer or the soft target. Each model was trained for one epoch (4,620 optimizer steps) with AdamW and a one-cycle schedule, at a peak learning rate of 3×10*^−^*^5^ for RadKev-27B and 2×10*^−^*^5^ for RadKev-9B and an effective batch size of 8 records, with bfloat16 computation on four NVIDIA RTX A6000 GPUs; the frozen weights were held in bfloat16 precision for RadKev-27B and in 32-bit precision for RadKev-9B. Both models were trained with data-parallel processes whose adapter and pointer-head gradients were summed before each optimizer step, which, because the loss is normalized by the number of records in the step, is equivalent to training in a single process: RadKev-27B with two processes, each holding the frozen backbone split across one NVLink-connected pair of GPUs and processing micro-batches of 2 records with 2 accumulation steps (13.1 hours), and RadKev-9B with four single-GPU processes and micro-batches of 2 records without accumulation (3.4 hours). A temperature was then fitted for each model on the development split by minimizing the negative log-likelihood [45] (1.35 for both models). Figure 3 shows the training loss, training accuracy and time per optimizer step of both models and their accuracy on the development split. Supplementary Note S3 gives the model revisions, the remaining hyperparameters and the software versions.

**Figure 2.**
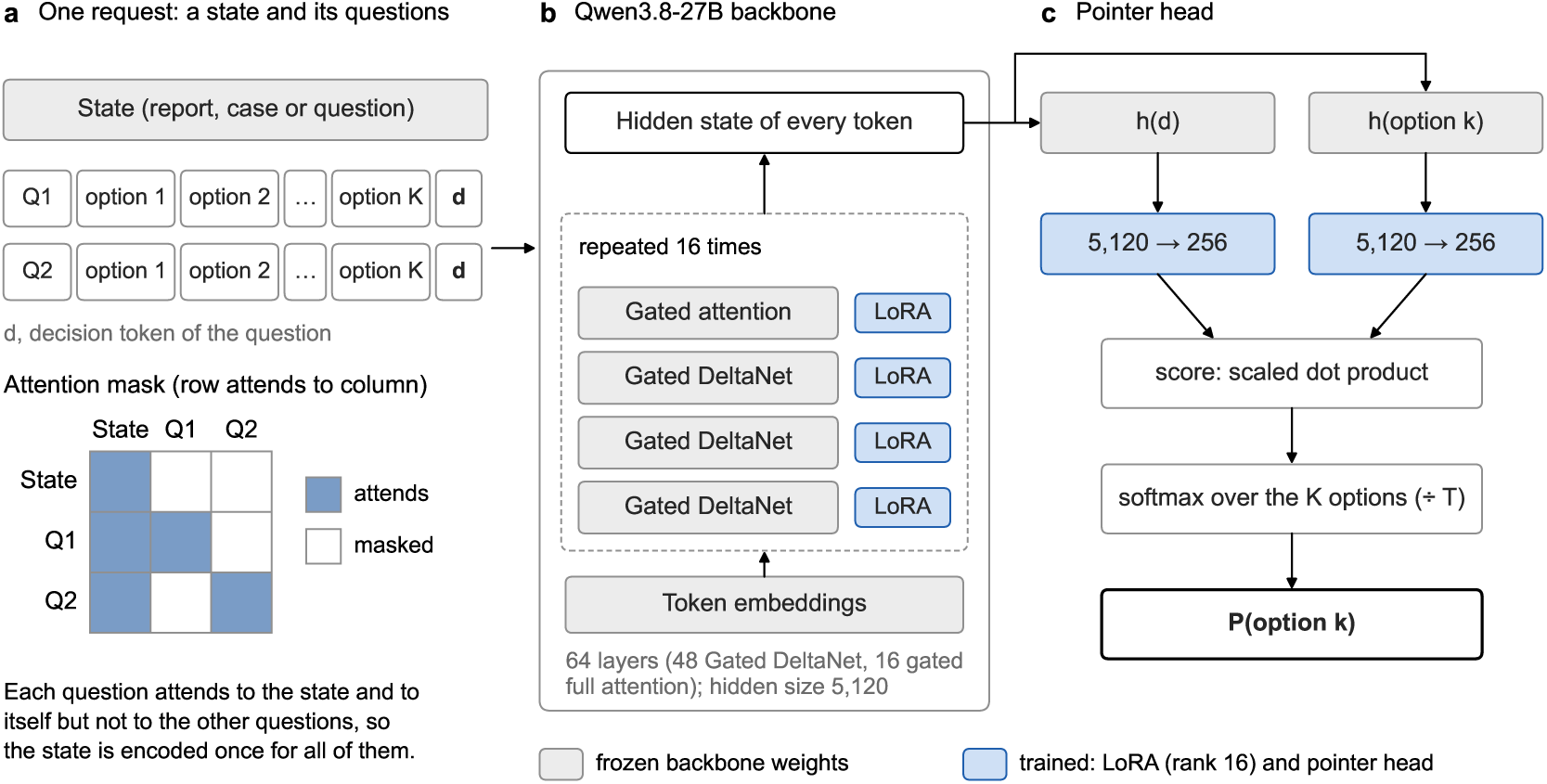
Architecture of Kev, the decision model fine-tuned in this study, shown for Kev-27B. (a) A request consists of a state followed by one or more questions, each with its options and a decision token; every question attends to the state and to itself but not to the other questions, so the state is encoded once. (b) The backbone is Qwen3.8-27B: 64 layers in 16 repeating groups of three Gated DeltaNet (linear-attention) layers and one gated full-attention layer, with hidden size 5,120. Its weights are frozen, and a rank-16 low-rank adapter (LoRA) is attached to every linear projection. (c) The pointer head projects the hidden states of the decision token and of each option’s closing token to 256 dimensions; the score of an option is the scaled dot product of the two projections, and the answer distribution is the softmax over the question’s options after division by a temperature T. Kev-9B has the same structure on Qwen3.5-9B-Base.

**Figure 3.**
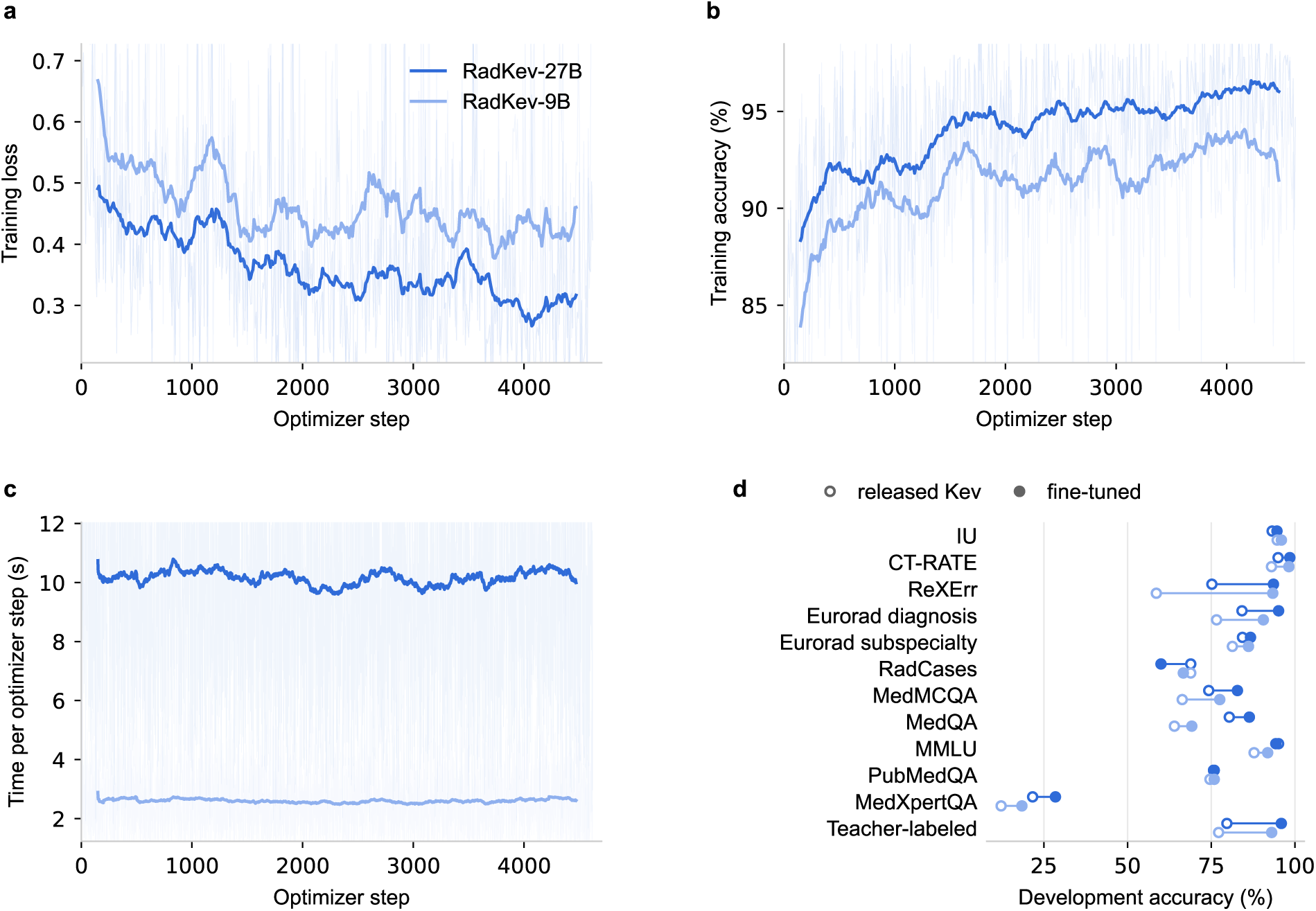
Training of RadKev-27B (dark blue) and RadKev-9B (light blue). (a) Training loss and (b) training accuracy on the micro-batches of the first data-parallel process, logged every 10 optimizer steps, and (c) wall-clock time of every optimizer step; thin lines show the logged values and thick lines a moving average over 300 optimizer steps, to whose range the vertical axes are scaled, so that logged values outside it are not shown. The loss is the cross-entropy over each question’s options on the radiology training records and the replayed Kev records, and the accuracy is the proportion of questions in the logged micro-batches answered correctly. (d) Accuracy on the development split, by source, of the released Kev models (open markers) and of the fine-tuned models (filled markers).

### 2.4 Evaluation

The models, the benchmark and the outcomes were specified in an addendum to the study’s analysis plan, written before any result of these models was read. The benchmark comprises 15 tasks: three IU/Open-i tasks (finding present, normal study and which finding), Eurorad diagnosis and subspecialty classification, the RadCases panel and topic questions, RSNA-RadioQA, ReXErr error detection, and one task for each examination source (the MedMCQA radiology subject, the other MedMCQA subjects, MedQA, MedXpertQA, MMLU and PubMedQA), in which that source’s radiology questions were pooled because the radiology selection left some subjects with few questions. The primary outcome was task-averaged accuracy, the unweighted mean of the accuracies of the 15 tasks, compared between RadKev-27B and Kev-27B. Secondary outcomes were the same mean over the 14 human-labeled tasks; accuracy over all benchmark questions and on every task; the differences between RadKev-27B and Qwen3.8-27B; expected calibration error and Brier score; the rate of confident errors; and coverage at a 5% error budget [1]. Every decision model was scored on the same test records, which contain the 14,142 benchmark questions, and received each record once, with all of its questions in a single request. Released models that are not pinned to a revision were loaded from fixed local snapshots, whose hashes were recorded. The comparators separate the effects of specialization, scale and model type: Kev-27B and Kev-9B [15], the starting points of RadKev-27B and RadKev-9B, measure specialization, and their difference measures scale within the same family; Qwen3.8-27B, the backbone of Kev-27B and RadKev-27B, measures the same network queried directly as an LLM; and the hosted general-purpose decision models Jev and the OpenAI Decisions API represent the externally hosted alternative (Section 2.6). The LLM was scored with the teacher prompt, one question per prompt, from the softmax of the next-token logits over the option letters with reasoning disabled, on every benchmark question with at most 16 options, which excludes the RadCases topic question with its 225 options, and every comparison with the LLM used the questions answered by both systems. Post hoc, the LLM was also scored with reasoning enabled on a sample of 1,675 benchmark questions (Supplementary Note S4).

The residual option of the RadCases panel question, stating that no ACR Appropriateness Criteria topic applies, was the most frequent panel answer in the training split (88 of 258 questions, 34%), and the specialized models reproduced this frequency as a prior and selected the option preferentially (Section 3.1). Their answer distributions on this question were therefore corrected post hoc for the training answer frequencies [46]: the probability of each of the 12 options was divided by that option’s relative frequency among the training-split panel answers, estimated with add-one smoothing, and the distribution was renormalized, which corresponds to assuming that every option is equally frequent at test time. The correction uses only training labels and involves no retraining. It was applied to RadKev-27B and RadKev-9B, which were trained on RadCases, and not to the other systems, which were not, and all results reported for RadKev-27B and RadKev-9B include it.

### 2.5 Statistical analysis

All comparisons were paired: both systems were evaluated on identical questions, and differences are reported as the first system minus the second. Ninety-five percent confidence intervals were obtained by a cluster bootstrap [47] with 2,000 resamples, in which test records were resampled within each source together with all of their questions, so that questions about the same report or case were never separated. One set of resamples was shared by all systems, so that every difference, and every difference between two differences, was computed on the same resamples. Intervals are the 2.5th and 97.5th percentiles of the resampled statistic; for task-averaged accuracy, the statistic is the mean of the per-task differences, and differences in calibration, confident errors and coverage were estimated on the first 800 resamples. Intervals for Kev’s transfer suite, which consists of different questions, were obtained with 1,000 resamples. No formal sample-size calculation was performed; the test set comprised all eligible held-out records of each source (Table 1). Two-sided *p* values were derived from the bootstrap distribution and adjusted by the Holm procedure [48] across the 15 tasks for the primary comparison, as specified in the addendum, and within each other set of comparisons reported together, for which the adjustment was not prespecified.

### 2.6 Additional analyses

The following analyses, some specified in the addendum and some added later, are exploratory; none involved training. The effect of specialization was compared with that of scale as a difference of differences, the gain from specialization (RadKev-27B minus Kev-27B, or RadKev-9B minus Kev-9B) minus the gain from scale (Kev-27B minus Kev-9B), computed on identical resamples. To test whether specialization reduced general decision performance, each specialized model and its starting checkpoint were compared on Kev’s out-of-domain transfer suite (656 questions). The gain of each specialized model was compared between withheld and seen instruction wordings; and calibration was re-estimated after recalibrating every system on the test split by two-fold cross-fitted temperature scaling.

An options-only control tested whether the models need the case to answer case-based questions. Every Eurorad diagnosis question of the test split and every RSNA-RadioQA question was posed again with the case removed: the state, which contains the patient’s age, sex, clinical history and imaging findings (Eurorad) or the case description (RSNA-RadioQA), was replaced by the single sentence “No case information is available.”, while the instructions and the answer options were left unchanged. A model that relies on the case should then fall to the accuracy expected by chance, one divided by the number of options; accuracy that remains well above chance indicates that the correct option can be recognized from the options themselves. RadKev-27B, Kev-27B, RadKev-9B, Kev-9B and Qwen3.8-27B were scored with and without the case, and the gains from specialization were compared between the two conditions. In addition, Eurorad diagnosis questions were divided according to whether a distinctive word of the correct option, at least six letters long and absent from the other options, appears in the case text.

Because a study is routed to a subspecialty reading list before its images are interpreted, the Eurorad subspecialty question was also posed with the imaging findings removed from its state and age, sex and clinical history retained (pre-read condition).

The effect of the answer space was examined on 30 Eurorad diagnosis questions of the test split and 30 RSNA-RadioQA questions by holding the case and the correct answer fixed and replacing the other options with randomly chosen or semantically similar diagnoses, for answer spaces of 2, 4, 16, 64 and 255 options, or with plausible incorrect answers generated by Qwen3.8-27B, for up to 16 options (Supplementary Note S5). Because single-letter scoring is limited by the number of letters, the LLM was scored on these answer spaces by greedy generation of the number of the chosen option, at every size, and additionally by option-letter probabilities up to 16 options; accuracy and latency were measured as a function of the number of options for every system. The hosted decision models, OpenAI Decisions and Jev, were scored on the same answer spaces, with each record sent once with all of its answer spaces as in the main evaluation, and timed end to end from the institution’s network on the same latency requests; questions that an API declined were counted as incorrect.

Latency, the measure of inference cost, was measured on 60 records drawn at random from the benchmark, on identical hardware, one request at a time, on one NVLink pair of RTX A6000 GPUs with Hugging Face Transformers in bfloat16; a decision model answered all questions of a record in one pass, whereas an LLM was called once per question, with option-letter scoring, with direct generation of the answer and, for Qwen3.8-27B on 55 questions (the first excluded as warm-up), with reasoning.

The OpenAI Decisions API [13] (model gpt-6-luna, which at the time of writing is the only model available through the API), an externally hosted general-purpose decision model released in public beta during the study, was scored on the same benchmark questions on 6 October 2026. Each benchmark record was sent once, as a POST request to the /v1/decisions endpoint whose JSON body contained three fields: model (gpt-6-luna); input, the state of the record; and questions, all benchmark questions of the record, each with a type, a name and the instructions that every other system received. Yes/no questions were sent as predicate questions; multiple-choice questions as choice questions, whose choices gave each option as a value (its identifier) and a description (its text); and ordinal questions as score questions with one entry in levels per grade. No other parameters were set and no additional prompting was used; CT-RATE questions were not sent. For each question, the API returns the probability of true (predicate) or a probability for each value (choice and score), which was scored as for the other systems; because these probabilities are rounded to two decimal places, the API was not recalibrated. Requests that failed with a rate-limit or server error were retried up to five times with exponential backoff, and questions that the API declined (5) received a uniform distribution. Latency was measured end to end, from the institution’s network to the response, for 27 requests sent one at a time and for the remaining 7,471 requests sent with four in flight, and the processing time reported by the API was recorded. The entire benchmarking run cost US$0.35.

Jev [11] (model jev-latest, which resolved to jev-1.13.0 in every response) was scored on the same benchmark questions in the same way on 7 October 2026. Because Jev accepts the request format of the benchmark, each record was sent once as a POST request to the /v1/systemone endpoint whose JSON body contained three fields: model; state, the state of the record; and questions, which mapped an anonymous name to each benchmark question of the record with its type, instructions and criteria exactly as every other system received them. No ground-truth label or other field was sent, no other parameters were set and CT-RATE questions were not sent. Jev returns the probability of true for yes/no questions and a probability for each option or grade otherwise; these were scored as for the other systems and, because they are rounded to two decimal places, were not recalibrated. Retries and latency measurement followed the protocol for the OpenAI Decisions API, except that Jev reports no processing time. The benchmarking run cost US$0.17.

As an external test on reports that no model was trained on, RadGraph-XL [21] provides 2,000 reports annotated by radiologists, 500 each of chest radiographs, chest CT, abdominal and pelvic CT and brain MRI. Reports that overlapped with a CheXpert Plus report or a RadKev record (a word 8-gram overlap of 50% or more) were excluded (495, all of them chest radiograph reports), leaving 1,505 reports. From each, up to three annotated observations were sampled, at most one per status, and posed as the four-option status question of the training format (present, explicitly absent, possible or equivocal, not mentioned), with the radiologists’ annotation as the ground truth (4,037 questions). Of these, 1,046 (25.9%) concerned hedged findings, which the report itself leaves open, as in “possible small hepatic cyst” or “early ischemia cannot be excluded”, and which the radiologists annotated as uncertain. Because their correct status depends on how the hedge is read rather than on what the report states, accuracy was analyzed on the 2,991 questions about definite findings, annotated as present or absent, with the uncertain option removed from the answer of every system and its probability renormalized over the remaining options; the hedged findings were analyzed separately, and the results on all 4,037 questions are given in Supplementary Table S12. RadKev-27B and RadKev-9B, their starting points, Qwen3.8-27B and the hosted decision models Jev and OpenAI Decisions were scored on this test set in the same way as on the benchmark (Supplementary Note S6).

## 3 Results

### 3.1 Primary comparison

Averaged over the fifteen benchmark tasks, the prespecified primary outcome, accuracy was 80.8% for RadKev-27B and 75.9% for Kev-27B, a difference of 4.9 percentage points (95% CI 3.8 to 6.2; Figure 4a). Over the fourteen tasks with human-labeled ground truth, the difference was 3.6 points (2.4 to 5.0), and over all 14,142 benchmark questions, each weighted equally, 5.8 points (5.3 to 6.3). The per-task differences ranged from 0.2 points (IU: finding present) to 22.6 points (ReXErr: report error), and after Holm adjustment across the fifteen tasks (Supplementary Table S2), RadKev-27B was more accurate on four tasks (IU: normal study, Eurorad: diagnosis, MedMCQA: other subjects, ReXErr: report error) and less accurate on no task.

**Figure 4.**
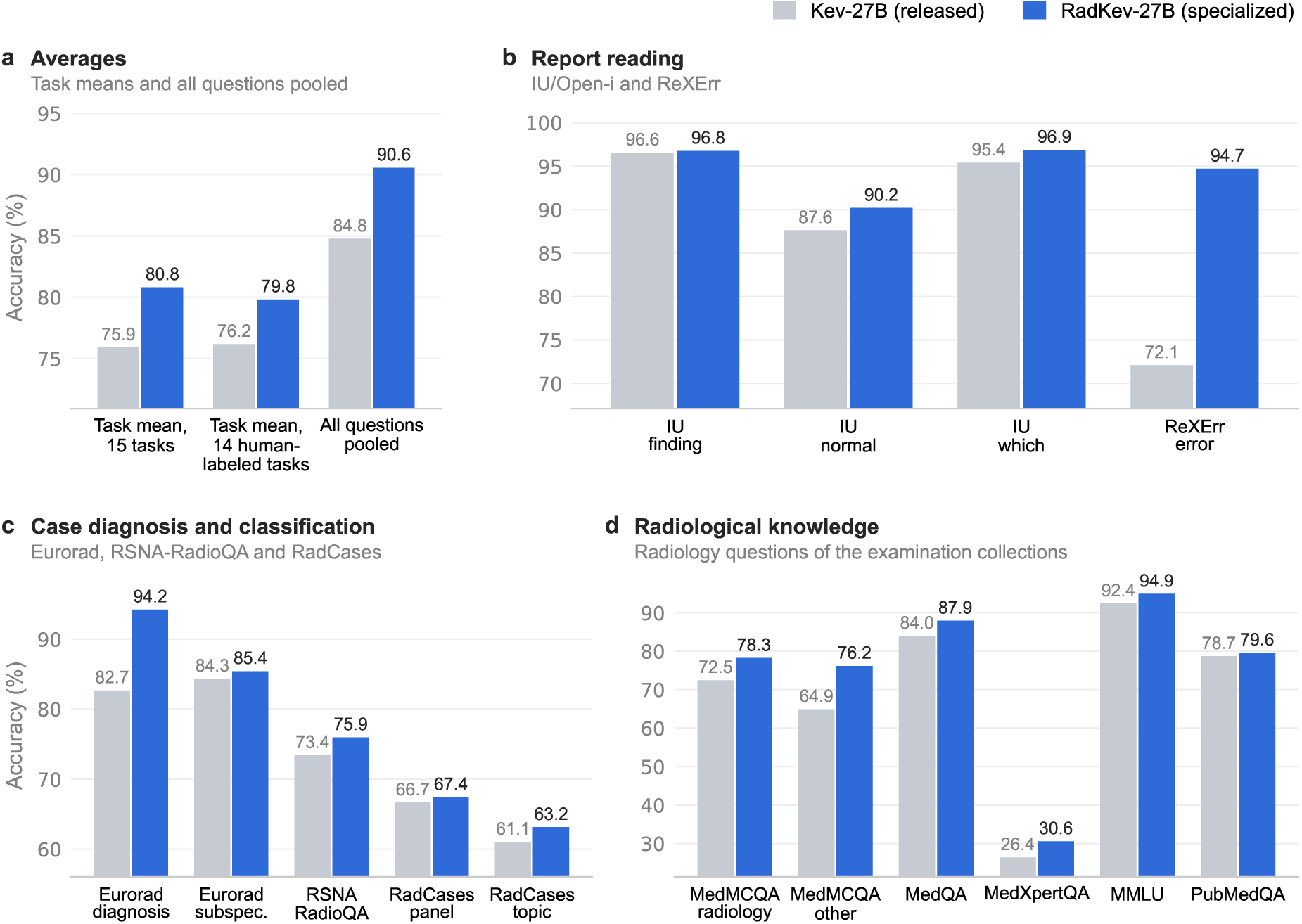
Accuracy of Kev-27B (grey) and of RadKev-27B (blue), the model specialized from it, on the radiology benchmark. (a) Averages: the unweighted mean of the task accuracies over all fifteen tasks (the primary outcome) and over the fourteen tasks with human-labeled ground truth, and accuracy over all 14,142 questions. (b) Report reading: IU/Open-i (finding present, normal study, which finding) and ReXErr (whether a report contains an error; ground truth known by construction). (c) Case diagnosis and classification: Eurorad diagnosis and subspecialty, RSNA-RadioQA, and the ACR Appropriateness Criteria panel and topic of RadCases. (d) Radiological knowledge: the radiology questions of the examination collections. Differences and 95% confidence intervals are given in the text and in Supplementary Table S2. Each accuracy axis spans 5 percentage points below the lowest and above the highest value in its panel.

In report reading (Figure 4b), the difference was 22.6 points (20.9 to 24.3) in the detection of errors in ReXErr reports, a task whose training split was part of the specialization data, and 0.2 (−0.1 to 0.5), 2.6 (1.6 to 3.5) and 1.5 points (0.3 to 2.7) on the IU/Open-i questions on finding presence, normal studies and which finding a report describes. In case diagnosis and classification (Figure 4c), the differences were 11.6 points for Eurorad diagnosis (8.1 to 15.6), 1.1 for Eurorad subspecialty classification (−2.2 to 4.5) and 2.5 for RSNA-RadioQA (79 questions; −2.5 to 7.6); on RadCases, whose training split was also used for specialization, they were 0.8 points for the ACR panel (−7.6 to 9.6) and 2.1 for the topic among 225 options (−2.0 to 6.6). On the radiology questions of the examination collections (Figure 4d), the mean of the six per-task differences was 4.8 points: 5.8 for MedMCQA radiology (−4.6 to 16.7), 11.3 for the radiology questions of the other MedMCQA subjects (5.7 to 17.3), 3.9 for MedQA (0.9 to 6.9), 4.2 for MedXpertQA (1.2 to 7.3), 2.5 for MMLU (0.0 to 6.3) and 0.9 for PubMedQA (−1.9 to 4.6).

The answer space of the RadCases panel question includes a residual option stating that no ACR Appropriateness Criteria topic applies, which was the correct answer to 88 of the 258 panel questions in the training split (34%), the most frequent answer. As scored, the specialized models selected this option preferentially: RadKev-27B selected it for 70.7% and RadKev-9B for 68.7% of the 99 questions whose correct answer was a panel (Kev-27B, 5.1%; Kev-9B, 5.1%), and their accuracy on the task was 43.9% and 44.7%. After the prior correction for the training answer frequencies (Section 2.4), they selected it for 11.1% and 21.2% of these questions and answered correctly 84.8% and 93.9% of the 33 questions for which it was the correct answer, and their accuracy was 67.4% and 68.2%, compared with 66.7% and 75.0% for the models they were specialized from (differences, 0.8 points, −7.6 to 9.6, and −6.8 points, −14.3 to 1.5). Qwen3.8-27B erred in the opposite direction and selected the residual option for 12.1% of the questions for which it was the correct answer. Without this correction, the difference in the primary outcome between RadKev-27B and Kev-27B was 3.3 points (2.1 to 4.8). Over the seven tasks from sources used only for evaluation (IU/Open-i, RSNA-RadioQA, MMLU, PubMedQA and MedXpertQA), the mean of the per-task differences was 2.1 points, and over the eight tasks from sources whose training splits were part of the specialization data, 7.4 points. Agreement with the labels of the CT-RATE classifier, which are not part of the benchmark, averaged 98.8% for RadKev-27B and 95.8% for Kev-27B over the three CT-RATE questions (Supplementary Table S3).

### 3.2 Decision models and language models

On the thirteen human-labeled tasks that every system answered, task-averaged accuracy was 81.1% for RadKev-27B, 79.4% for OpenAI Decisions, 79.1% for Jev, 78.1% for Qwen3.8-27B, 77.3% for Kev-27B, 77.0% for RadKev-9B and 72.0% for Kev-9B (Figure 5a; Supplementary Table S3). On the questions with at most 16 options (fourteen tasks), RadKev-27B was more accurate than Qwen3.8-27B, its own backbone queried as an LLM (task mean, 4.7 points, 95% CI 3.4 to 6.0; over the thirteen human-labeled tasks, 3.0 points, 1.6 to 4.5; Figure 5b; Supplementary Table S4). The differences from Qwen3.8-27B ranged by task from −3.4 points (IU: normal study) to 25.6 points (ReXErr: report error) and were positive on ten of fourteen tasks. Before specialization, Qwen3.8-27B was not detectably different in accuracy from Kev-27B (0.4 points, −0.8 to 1.7). RadKev-9B was not detectably different in accuracy from Qwen3.8-27B (0.7 points, −1.0 to 2.5). OpenAI Decisions (gpt-6-luna), a hosted general-purpose decision model, reached a benchmark task mean of 76.0%. RadKev-27B was more accurate than it (4.8 points, 3.2 to 6.4; over the human-labeled tasks, 4.0 points, 2.3 to 5.7), with per-task differences from −10.0 points (MedXpertQA) to 33.7 points (RadCases: ACR topic), and RadKev-9B was not detectably different in accuracy from it (1.0 points, −0.8 to 2.9). The API declined 5 of the 14,142 questions, which were scored as a uniform distribution over their options. Measured end to end from the institution’s network, its median latency per question was 145 ms with four requests in flight (7,471 requests) and 175 ms with one request at a time (27 requests), and the processing time reported by the API was 45 ms per request (median). Jev (jev-1.13.0), the hosted general-purpose decision model whose interface Kev reproduces, reached a benchmark task mean of 77.8% and returned a probability distribution for every question. It was more accurate than Kev-27B (1.9 points, 0.3 to 3.4) and not detectably different in accuracy from Qwen3.8-27B (1.4 points, 0.0 to 2.8), and OpenAI Decisions was less accurate than it (−1.8 points, −3.4 to −0.3). RadKev-27B was more accurate than Jev (3.0 points, 1.7 to 4.4; over the human-labeled tasks, 1.9 points, 0.4 to 3.3), with per-task differences from −2.5 points (MMLU) to 19.1 points (ReXErr: report error), and RadKev-9B was not detectably different in accuracy from it over all tasks (−0.8 points, −2.5 to 0.9) and less accurate than it over the human-labeled tasks (−2.1 points, −3.8 to −0.3). Measured end to end from the institution’s network, the median latency of Jev per question was 168 ms with four requests in flight (7,471 requests) and 185 ms with one request at a time (27 requests). In the post hoc analysis with reasoning enabled, on a sample of 1,675 benchmark questions on which every system was scored, Qwen3.8-27B reached 83.6% without and 85.1% with reasoning (1.6 points, 0.1 to 3.0); RadKev-27B, which does not reason, reached 89.0%, a difference from Qwen3.8-27B with reasoning of 3.8 points pooled over questions (2.2 to 5.5) and 2.8 points averaged over the nine tasks of the sample, with the examination questions pooled into one task (1.0 to 4.6; Supplementary Table S5 and Supplementary Note S4). On 60 benchmark records timed one request at a time, the median latency per question was 128 ms for RadKev-27B and 43 ms for RadKev-9B, compared with 145 ms for Qwen3.8-27B scored from its option-letter logits, 234 ms when it generated the answer letter and 16.9 s with reasoning, approximately 132 times the latency of RadKev-27B (Figure 5c; Supplementary Table S6).

**Figure 5.**
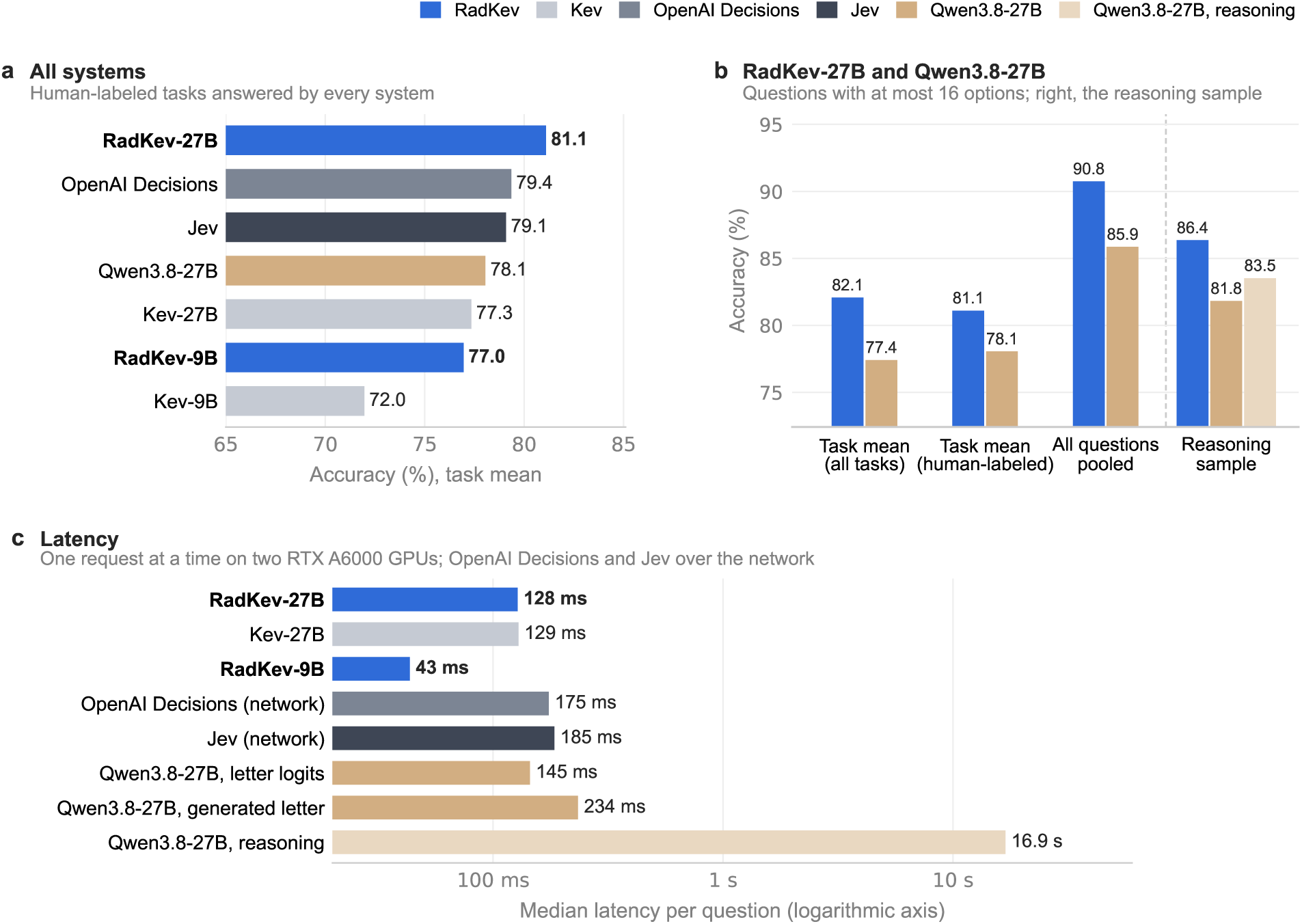
Decision models and language models. (a) Task-averaged accuracy of every system over the thirteen human-labeled tasks that all systems answered (Qwen3.8-27B was not scored on the RadCases topic question, which has 225 options). (b) RadKev-27B and Qwen3.8-27B on the benchmark questions with at most 16 options, averaged over tasks and pooled over questions, and, at right, on the reasoning sample of 1,675 questions, averaged over its nine tasks, with Qwen3.8-27B also scored with reasoning enabled. The accuracy axes span 5 percentage points below the lowest and above the highest value. (c) Median latency per question, one request at a time on two RTX A6000 GPUs (logarithmic axis); a decision model answers all questions about a record in one pass, Qwen3.8-27B was scored from its option-letter logits, by generating the answer letter, or with reasoning, and the hosted OpenAI Decisions API and Jev were timed end to end from the institution’s network, one request at a time (27 requests each). Blue, RadKev; grey, the other decision models (Kev, light; OpenAI Decisions, medium; Jev, dark); tan, Qwen3.8-27B (with reasoning, light).

### 3.3 Specialization and scale

Specialization changed the benchmark task mean by 4.9 points at 27B and by 6.9 points at 9B (5.5 to 8.4), and the threefold increase in size from Kev-9B to Kev-27B changed it by 5.8 points (4.2 to 7.5; Figure 6a); the difference between the two gains (specialization minus scale) was −0.9 points at 27B (−3.1 to 1.6) and 1.1 points at 9B (−0.5 to 2.7); the latter equals the difference between RadKev-9B and Kev-27B, so the specialized 9B model was not detectably different in accuracy from the general-purpose model three times its size. Over the human-labeled tasks, the gain from specialization at 27B (3.6 points) was not detectably different from that from scale (5.1 points; difference −1.5, −3.9 to 1.1), whereas pooled over the human-labeled questions, of which IU/Open-i report reading forms the majority, the corresponding gains were 1.8 and 0.3 points (difference 1.5, 0.8 to 2.2; Figure 6b), so the comparison of specialization with scale depends on how tasks are weighted. On Kev’s out-of-domain transfer suite (656 questions), which measures general decision skill outside radiology, accuracy changed by 1.5 points for RadKev-9B relative to Kev-9B (0.0 to 3.1) and by −0.6 points for RadKev-27B relative to Kev-27B (−2.3 to 1.2; Figure 6c; Supplementary Table S7).

**Figure 6.**
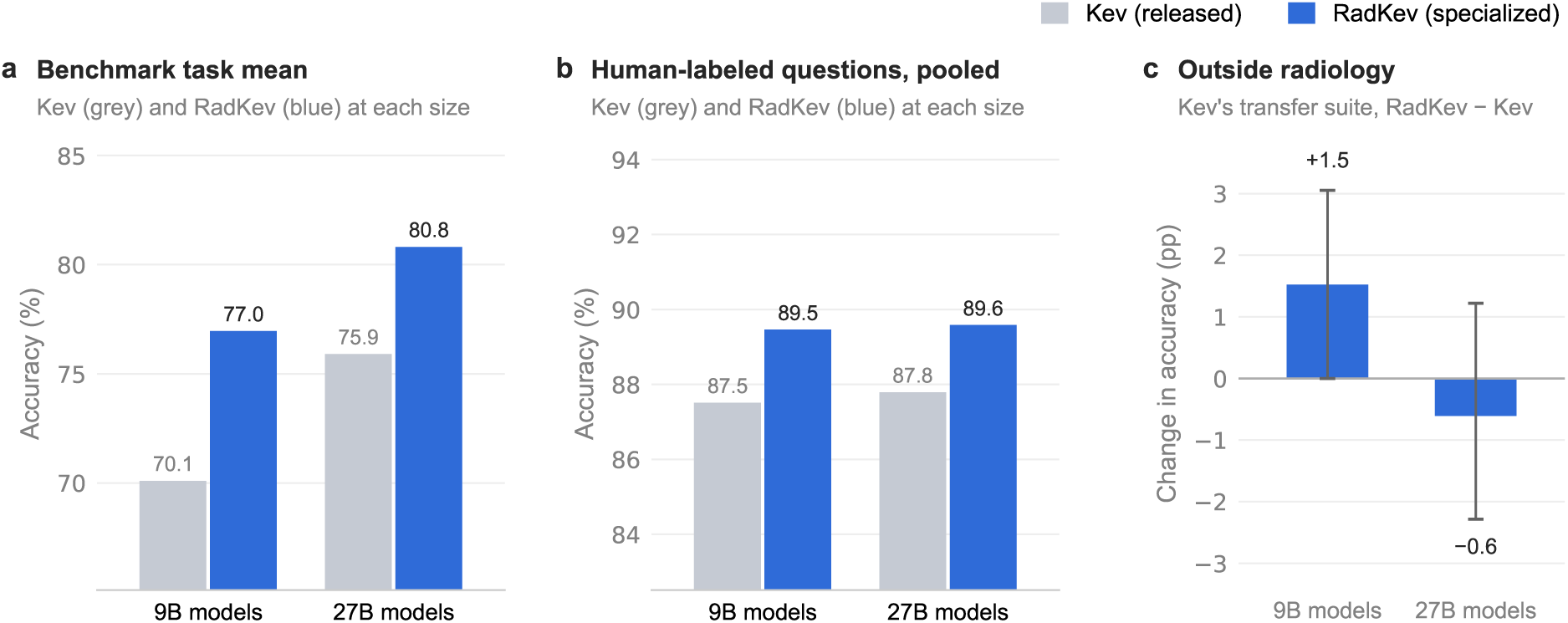
Specialization and scale. The released Kev models (grey) and their specialized counterparts, RadKev (blue), at 9B and 27B, as the benchmark task mean (a) and pooled over the human-labeled questions (b); the gain from specialization is the difference within each pair, and the gain from scale is the difference between the two grey bars. Each accuracy axis spans 5 percentage points below the lowest and above the highest value. (c) Change in accuracy on Kev’s out-of-domain transfer suite, specialized minus released model; whiskers, 95% confidence intervals.

### 3.4 Calibration and selective prediction

On the human-labeled questions, answered in descending order of confidence, RadKev-27B could answer 88.4% of them with an error rate of at most 5% among the answered questions, compared with 85.5% for Kev-27B (2.9 points, 95% CI 1.9 to 3.9), 86.6% for Qwen3.8-27B, 85.9% for OpenAI Decisions and 86.8% for Jev (RadKev-27B minus OpenAI Decisions, 2.5 points, 0.8 to 4.6; minus Jev, 1.6 points, 0.6 to 2.8; Figure 7a). Over all 14,142 benchmark questions, including the ReXErr questions, the corresponding coverage was 90.5% for RadKev-27B and 76.4% for Kev-27B. The expected calibration error of RadKev-27B was 0.015, compared with 0.018 for Kev-27B and 0.022 for Qwen3.8-27B; its Brier score was 0.149 (Kev-27B, 0.164); and 1.4% of questions were answered incorrectly with a confidence of at least 0.9, compared with 0.6% for Kev-27B (Figure 7b, c). After cross-fitted recalibration on the test split, the calibration errors of RadKev-27B and Kev-27B were 0.015 and 0.015, and their coverage at a 5% error budget 88.2% and 85.3% (Supplementary Table S8).

**Figure 7.**
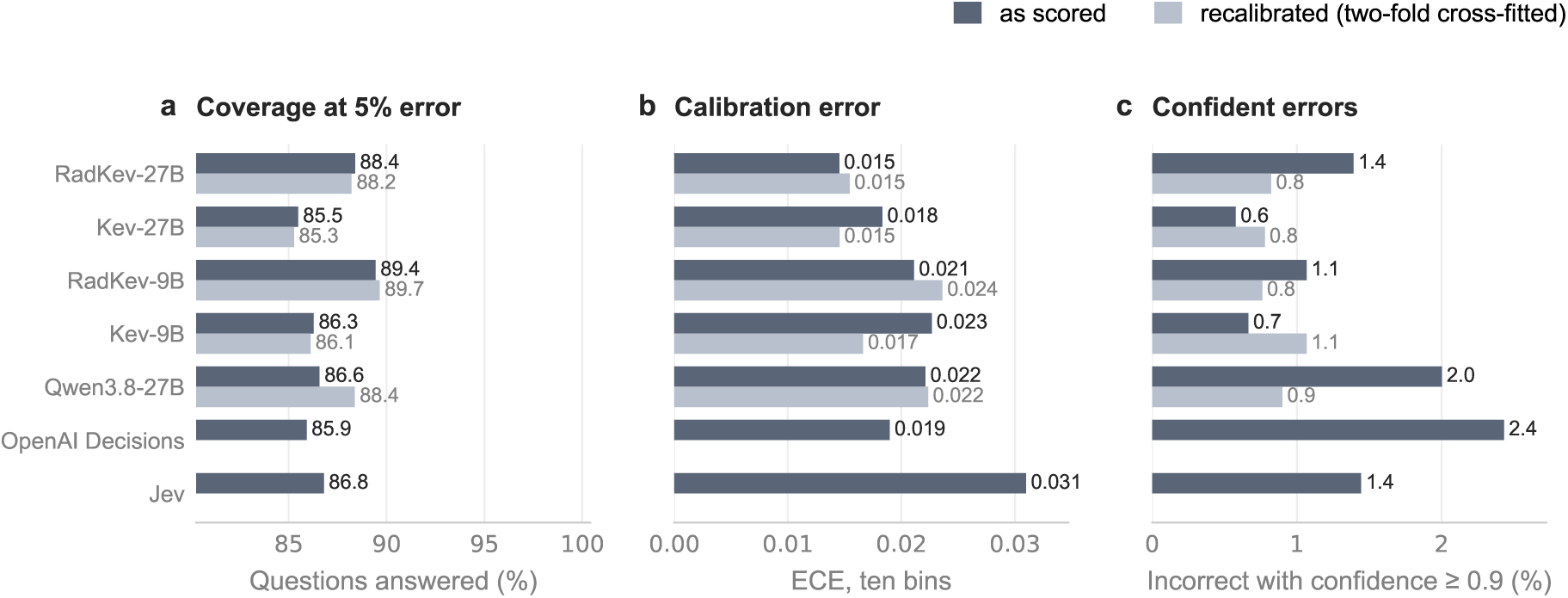
Calibration and selective prediction on the 11,434 human-labeled benchmark questions. (a) Proportion of questions that can be answered, in descending order of confidence, with at most 5% error among the answered questions. (b) Expected calibration error over ten equal-width confidence bins. (c) Questions answered incorrectly with a confidence of at least 0.9. Dark bars show the models as scored, each with its own temperature, and light bars after two-fold cross-fitted temperature recalibration on the test split, which leaves accuracy unchanged. The hosted decision models, OpenAI Decisions and Jev, return probabilities rounded to two decimals and are shown as scored only.

### 3.5 Robustness of the gains

When the Eurorad subspecialty question was posed with the imaging findings removed, leaving the age, sex and clinical history that are available before a study is read, the accuracy of every system fell, by 11.9 to 17.8 percentage points (185 questions; Figure 8a; Supplementary Table S9). RadKev-27B reached 73.0% before the read and 85.4% with the findings; before the read, its difference from Kev-27B was 2.7 points (−1.6 to 7.0) and that from Qwen3.8-27B 2.7 points (−2.2 to 8.1). On the IU/Open-i, Eurorad and examination questions, the gain of RadKev-27B over Kev-27B was 0.7 points on instruction wordings withheld from training (0.1 to 1.2; 5,629 questions) and 2.9 points on wordings seen in training (2.3 to 3.6; 5,499 questions), a difference of −2.3 points (−3.1 to −1.5); for RadKev-9B, the corresponding gains were 2.1 (1.6 to 2.7) and 1.9 points (1.3 to 2.5), a difference of 0.2 points (−0.6 to 1.0; Figure 8b). In the options-only control, in which the case was removed and only the question and its answer options were shown, the accuracy of RadKev-27B on Eurorad diagnosis fell from 94.2% to 76.9%, that of Kev-27B from 82.7% to 47.7% and that of Qwen3.8-27B from 84.1% to 50.0%, against 24% expected by chance (Figure 8c). The gain from specialization was larger without the case (29.2 points, 24.0 to 34.7) than with it (11.6 points, 8.1 to 15.6; difference −17.6, −24.0 to −11.3), and at 9B it was 43.1 points without and 15.3 points with the case. With the case available, the gain was 7.3 points (3.7 to 11.2) in the 178 questions in which a distinctive word of the correct option occurred in the case text and 16.1 points (9.8 to 22.5) in the 168 in which none did, so it did not depend on words of the correct diagnosis appearing in the case. The specialized models therefore recognized the case author’s final diagnosis among the differential diagnoses from the answer options themselves, and the gain on Eurorad diagnosis does not indicate improved reading of the case. On RSNA-RadioQA, whose incorrect options were written by LLMs, options-only accuracy was 50.6% for RadKev-27B, 40.5% for Kev-27B and 46.8% for Qwen3.8-27B, against 25% expected by chance; the gain from specialization was 10.1 points without the case (−1.3 to 21.5) and 2.5 points with it (−2.5 to 7.6; Figure 8d; Supplementary Table S10).

**Figure 8.**
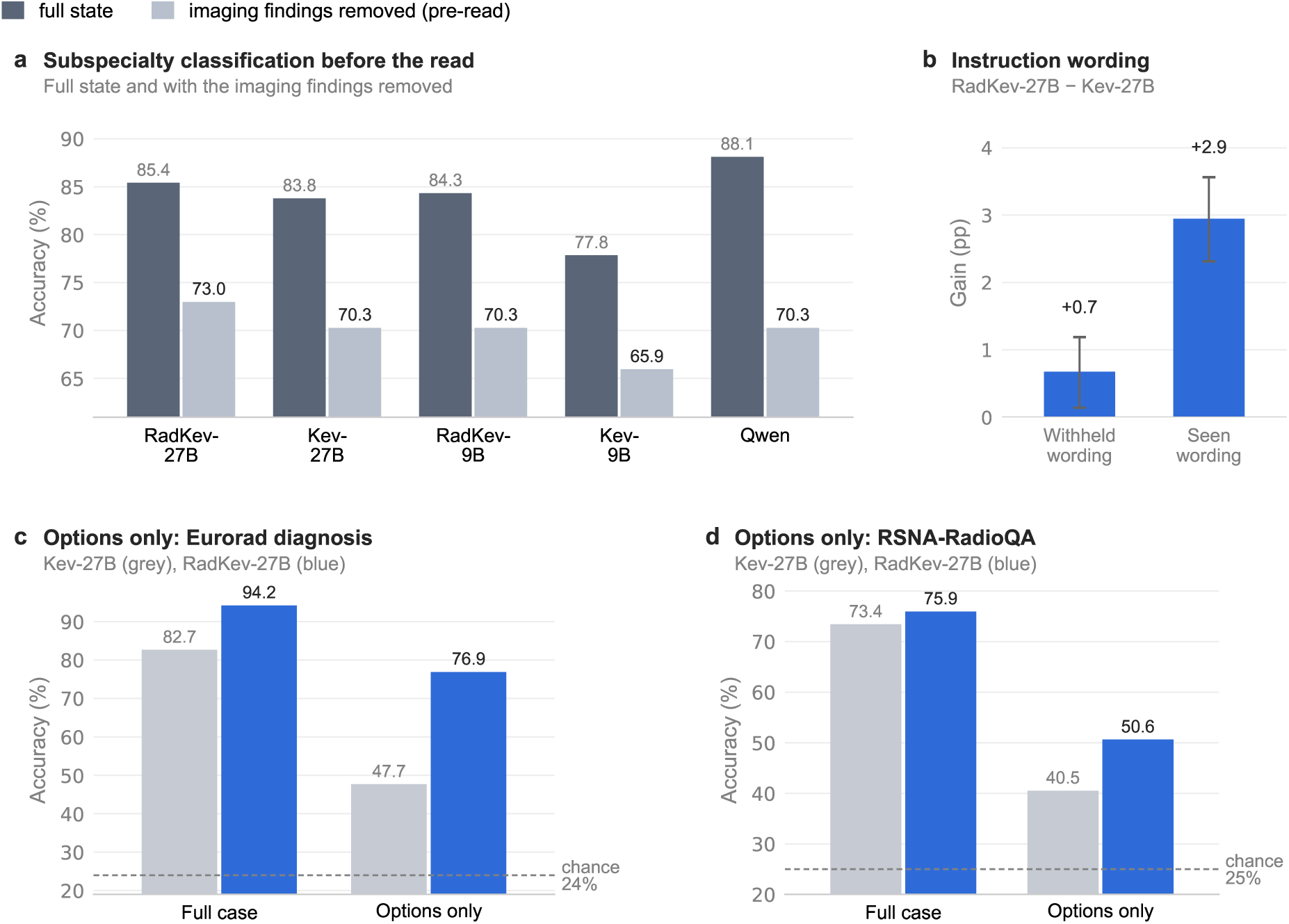
Robustness of the gains. (a) Eurorad subspecialty classification with the full state (dark) and with the imaging findings removed (light), as before the read; accuracies as recomputed in this analysis (Supplementary Table S9). (b) Gain of RadKev-27B over Kev-27B on instruction wordings withheld from and seen in training; whiskers, 95% confidence intervals. (c, d) Options-only control: accuracy of Kev-27B (grey) and RadKev-27B (blue) with the full case and with only the question and its options, for Eurorad diagnosis (c) and RSNA-RadioQA (d); a model that needs the case should fall to chance (dashed lines).

### 3.6 Size and composition of the answer space

In the answer-space analysis of 30 Eurorad diagnosis and 30 RSNA-RadioQA questions, in which the case and the correct answer were held fixed and the other options replaced by 1 to 254 alternatives, accuracy depended more on how similar the alternatives were to the correct answer than on how many were offered (Supplementary Table S11). With the 16 most similar Eurorad diagnoses as options, the accuracy of RadKev-27B was 93.3%, compared with 90.0% for Kev-27B and 83.3% for Qwen3.8-27B (Figure 9); with 255 options, it was 93.3% with random and 73.3% with the most similar alternatives. On RSNA-RadioQA, with the 16 most similar alternatives, the accuracy of RadKev-27B was 60.0%, compared with 63.3% for Kev-27B and 43.3% for Qwen3.8-27B; with 255 options, it was 83.3% with random and 33.3% with the most similar alternatives. Alternatives written by Qwen3.8-27B were less confusable than the most similar diagnoses of the pool: with 16 LLM-written options, RadKev-27B answered 100.0% of 18 Eurorad and 81.0% of 21 RSNA-RadioQA questions correctly. Pooled over both sources, the mean confidence of RadKev-27B exceeded its accuracy by 17.8 points with the 255 most similar alternatives (71.1% vs 53.3%); answers given with a confidence of at least 0.9 were correct in 92% to 100% of questions under every answer space, but the proportion of questions answered with that confidence fell from 93% with 16 random to 30% with 255 most similar alternatives. Qwen3.8-27B was scored by generating the number of the chosen option and, with up to 16 options, also from the probabilities of single-letter option labels; the decision models score each option directly. On 15 Eurorad cases with one question per request, the latency of RadKev-27B rose from 255 ms with two options to 3,414 ms with 255. The latency of Qwen3.8-27B rose from 331 ms with two options to 2,207 ms with 255 when it generated the number of its chosen option, and was 242 to 299 ms for 2 to 16 options when scored from the probabilities of the option letters. The hosted decision models were scored on the same answer spaces. With the 16 most similar alternatives, OpenAI Decisions and Jev were correct in 90.0% and 83.3% of Eurorad and in 56.7% and 56.7% of RSNA-RadioQA questions. With 255 options, OpenAI Decisions was correct in 60.0% of Eurorad questions with random and in 46.7% with the most similar alternatives (RSNA-RadioQA, 60.0% and 23.3%), and Jev in 96.7% and 80.0% (RSNA-RadioQA, 73.3% and 46.7%). OpenAI Decisions declined 3 of the answer-space questions. Measured end to end from the institution’s network, the latency per request of the hosted decision models changed little with the number of options: 175 ms with two and 216 ms with 255 options for OpenAI Decisions, and 184 and 212 ms for Jev.

**Figure 9.**
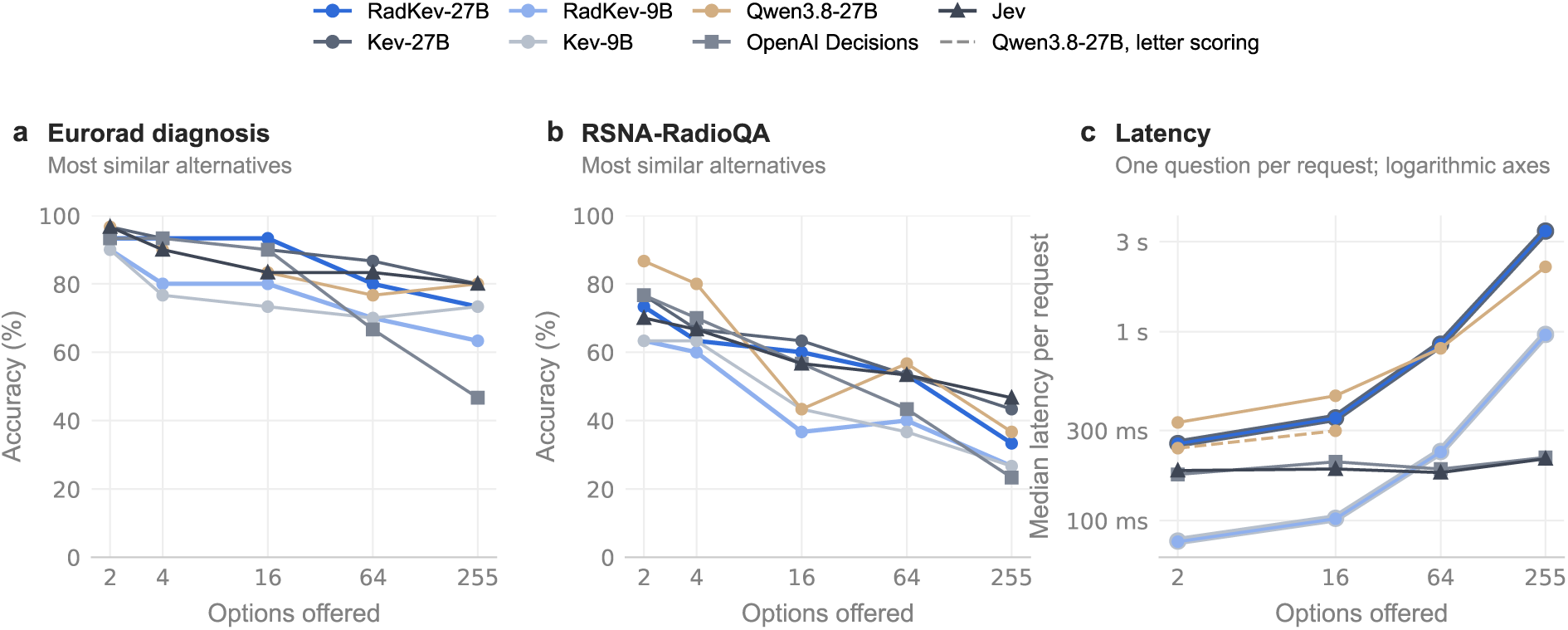
Accuracy and latency as the answer space grows (2, 4, 16, 64 and 255 options): 30 Eurorad diagnosis and 30 RSNA-RadioQA questions with the correct answer offered together with the most similar alternatives, and median latency per request with one question per request on 15 Eurorad cases, in which the lines of RadKev and Kev models of the same size coincide; the hosted decision models were timed end to end from the institution’s network. Qwen3.8-27B was scored by generating the number of the chosen option. Results with random and LLM-written alternatives are given in Supplementary Table S11; Supplementary Note S5 describes the construction of the answer spaces.

### 3.7 External test set

Of the 4,037 RadGraph-XL status questions, from predominantly CT and MRI reports of another institution, 2,991 concerned definite findings. On these, accuracy was 90.2% for RadKev-27B, 91.3% for Kev-27B, 90.3% for Qwen3.8-27B, 89.3% for Jev and 81.9% for OpenAI Decisions. Specialization changed the accuracy of the 27B model by −1.1 points (95% CI −1.6 to −0.6) and that of the 9B model by 2.4 points (1.5 to 3.3), and RadKev-27B was not detectably different in accuracy from Qwen3.8-27B (−0.1 points, −0.8 to 0.7), more accurate than Jev (0.9, 0.3 to 1.6) and more accurate than OpenAI Decisions (8.3, 7.3 to 9.3; Supplementary Table S12). On the 1,046 hedged findings, accuracy was 39.1% for RadKev-27B, 40.7% for Kev-27B, 55.4% for Qwen3.8-27B, 43.1% for Jev and 36.8% for OpenAI Decisions. Every decision model, including the two hosted ones, most often classified them as present, and they accounted for 174 of the 229 questions that only Qwen3.8-27B answered correctly; uncertain findings formed 6.7% of the status answers in the training data. Over all 4,037 questions, accuracy was 76.3% for RadKev-27B, 77.4% for Kev-27B and 78.4% for Qwen3.8-27B.

## 4 Discussion

Specializing a general-purpose decision model on public radiology data improved its task-averaged accuracy on a radiology benchmark by 4.9 percentage points, and the specialized 27B model was more accurate than its starting point, the hosted decision models Jev and OpenAI Decisions and its own backbone used as an LLM, over the benchmark as a whole and over the human-labeled tasks alone. On a sample of 1,675 questions, it was also more accurate than that backbone with reasoning enabled, at 132-fold lower latency. Before specialization, the backbone was approximately as accurate when queried as a general-purpose decision model as when queried as an LLM, so the advantage arose from specialization rather than from the decision format alone. Kev-27B, which reproduces the interface of Jev with open weights, was less accurate than Jev (1.9 points), and specialization reversed this ordering. At 9B, specialization raised the task mean by 6.9 points, and the specialized model did not differ detectably from a general-purpose model three times its size; at 27B, the gain from specialization did not differ detectably from that from a threefold increase in model size when tasks were weighted equally, and exceeded it when the human-labeled questions were pooled. Analyses other than the primary comparison are exploratory.

The gains were concentrated in a few tasks: they were largest in the detection of errors in reports and in Eurorad diagnosis, absent in finding presence and subspecialty classification, and larger on tasks from sources whose training splits were used for specialization (7.4 points) than on tasks from sources used only for evaluation (2.1 points). The options-only control qualifies the Eurorad gain: without the case, the specialized models still selected the correct diagnosis in most questions, and their gain over the general-purpose models was larger than with the case, so this gain reflects recognition of the case author’s final diagnosis among the options rather than reading of the case. The same caution applies to other benchmarks whose incorrect options are written by the authors of the correct answer. For RadKev-27B, part of the gain also depended on familiar question phrasing (0.7 points on withheld and 2.9 on seen instruction wordings), whereas for RadKev-9B it did not (2.1 and 1.9 points). Removing the imaging findings reduced the accuracy of every system on the subspecialty question, so classifying an interpreted case, as posed in the benchmark, is less demanding than routing a study before it is read.

Two results concern the answers in the training data. On the RadCases panel question, the residual option that no topic applies was the most frequent training answer (34%), and the specialized models selected it preferentially (accuracy 43.9% for RadKev-27B) until their answer distributions were corrected for the training answer frequencies (67.4%, against 66.7% for Kev-27B). On the definite findings of radiologist-annotated reports from another institution (RadGraph-XL), specialization changed the accuracy of the 27B model by −1.1 points (−1.6 to −0.6) and that of the 9B model by 2.4 points (1.5 to 3.3), and RadKev-27B was not detectably different in accuracy from its backbone used as an LLM (−0.1 points). The hedged findings were the principal source of error: before and after specialization, the open decision models, like the hosted ones, frequently classified them as present (correct in 40.7% for Kev-27B and 39.1% for RadKev-27B, against 55.4% for Qwen3.8-27B); such findings formed 25.9% of these questions but only 6.7% of the status answers in the training data, and specialization did not correct this tendency. The gains on the benchmark therefore do not carry over unchanged to reports from another institution, and the training answers should cover the answer distribution of the intended use.

Beyond accuracy, a decision model returns a probability for every admissible answer of every question in one pass, which permits automation of confident decisions and referral of uncertain ones [1]: at a 5% error budget, RadKev-27B could answer 88.4% of the human-labeled questions, compared with 85.5% for Kev-27B and 86.6% for Qwen3.8-27B. Its expected calibration error was similar to that of Kev-27B, but it made more confident errors (1.4% vs 0.6% of questions), which recalibration reduced (Section 4.1). Per question, an LLM scored from its option-letter logits was approximately as fast as the decision model (145 vs 128 ms), and with 255 options an LLM generating the number of its choice was faster (2,207 vs 3,414 ms); the latency advantage applies to LLMs that generate their answers, particularly with reasoning (16.9 s), and to records with several questions, which a decision model answers in one pass.

These results agree with earlier work on domain adaptation, in which continued training on domain text improved downstream performance [18] and lightweight adaptation of open models has been proposed for clinical and radiology text [49–52], although prompting alone has also allowed a generalist model to exceed medically fine-tuned models [19]. For decision models, which have been less accurate than frontier LLMs on medical questions (Section 1.3), the present results show that specialization can make an open model more accurate on radiology decisions than an LLM of the same size and than hosted general-purpose decision models. LLMs have been evaluated for the detection of errors in reports [4] and for selecting imaging under the ACR Appropriateness Criteria [5, 6]; in the present study, specialization on 6,000 labeled reports improved error detection by 22.6 points, whereas the assignment of ACR panels, with 264 training cases, did not differ detectably from that of the starting model.

### 4.1 Clinical implementation

#### Decisions and infrastructure

Table 2 summarizes the recommendations below and the results on which they rest; none of these uses was evaluated prospectively. A decision model suits frequent decisions on radiology text whose admissible answers can be listed in advance, such as examination selection, prioritization, subspecialty routing and the detection of critical findings or report errors [51, 53]. Each decision should be validated on the information available when it is made: removing the imaging findings reduced subspecialty accuracy by 11.9 to 17.8 points. The 27B model runs on two 48 GB GPUs and the 9B model on one (task mean 80.8% vs 77.0%), specialization took 13.1 h and 3.4 h on four such GPUs, and all decisions about a record should be posed in one request; unlike with a hosted decision model, patient data need not leave the institution.

**Table 2.**
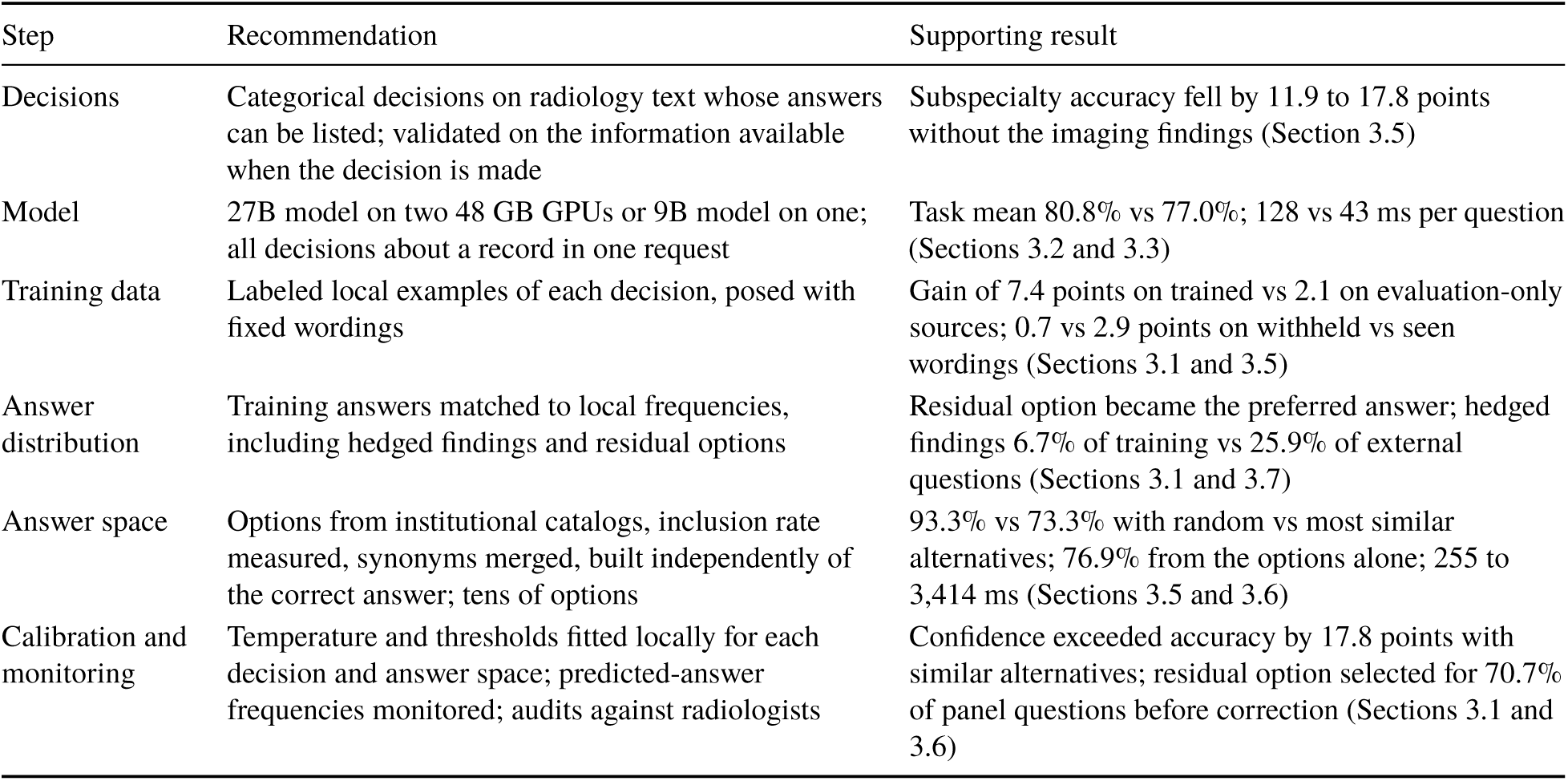
Recommendations for the clinical implementation of a specialized decision model and the results on which they rest.

| Step | Recommendation | Supporting result |
| --- | --- | --- |
| Decisions | Categorical decisions on radiology text whose answers can be listed; validated on the information available when the decision is made | Subspecialty accuracy fell by 11.9 to 17.8 points without the imaging findings (Section 3.5) |
| Model | 27B model on two 48 GB GPUs or 9B model on one; all decisions about a record in one request | Task mean 80.8% vs 77.0%; 128 vs 43 ms per question (Sections 3.2 and 3.3) |
| Training data | Labeled local examples of each decision, posed with fixed wordings | Gain of 7.4 points on trained vs 2.1 on evaluation-only sources; 0.7 vs 2.9 points on withheld vs seen wordings (Sections 3.1 and 3.5) |
| Answer distribution | Training answers matched to local frequencies, including hedged findings and residual options | Residual option became the preferred answer; hedged findings 6.7% of training vs 25.9% of external questions (Sections 3.1 and 3.7) |
| Answer space | Options from institutional catalogs, inclusion rate measured, synonyms merged, built independently of the correct answer; tens of options | 93.3% vs 73.3% with random vs most similar alternatives; 76.9% from the options alone; 255 to 3,414 ms (Sections 3.5 and 3.6) |
| Calibration and monitoring | Temperature and thresholds fitted locally for each decision and answer space; predicted-answer frequencies monitored; audits against radiologists | Confidence exceeded accuracy by 17.8 points with similar alternatives; residual option selected for 70.7% of panel questions before correction (Sections 3.1 and 3.6) |

#### Training data

The gains were largest where the training data contained the target decision (7.4 vs 2.1 points), so the model should be specialized on labeled local examples of each decision, with answer frequencies that match local use: the residual option, 34% of the RadCases training answers, became the preferred answer, and hedged findings, 6.7% of the training status answers but 25.9% of the external questions, remained the principal source of error. Questions should be posed with the wordings used in training (gain 0.7 vs 2.9 points on withheld vs seen wordings).

#### Answer spaces

The model can choose only among the options offered, so the inclusion rate, the proportion of local cases whose correct answer is offered, should be measured first. Options can be taken from institutional catalogs, triage scales or reporting templates or, for open decisions such as differential diagnosis, from a curated catalog or an LLM. The similarity of the alternatives mattered more than their number (93.3% vs 73.3% with 255 random vs most similar alternatives), so options should be mutually exclusive, synonyms merged and candidate lists validated in the form used. Options written together with the correct answer can reveal it (76.9% accuracy from the options alone), so answer spaces should be built independently of the correct answer and checked with an options-only control. The latency of RadKev-27B rose from 255 to 3,414 ms between 2 and 255 options, whereas that of the hosted decision models changed little, so a locally run model is best used with lists of tens of options.

#### Calibration and monitoring

Temperatures and confidence thresholds should be fitted on local data for each decision and answer space: with highly similar alternatives, mean confidence exceeded accuracy by 17.8 points, although answers given with a confidence of at least 0.9 remained correct in at least 92% of questions. At an error rate of 5%, 88.4% of the human-labeled questions could be automated and the remainder referred to a radiologist. In operation, predicted-answer frequencies should be compared with local expectations, which would have revealed the preference for the residual option (70.7% of panel questions) without labels, alongside periodic audits against radiologists.

### 4.2 Limitations

The findings should be interpreted within the scope of the study design. The evaluation used public data: the test splits of Eurorad, RadCases, ReXErr, MedMCQA and MedQA derive from the same collections as their training splits, the errors in ReXErr reports were introduced by an LLM, and generalization to reports from another institution was assessed on one external test set (RadGraph-XL). Validation on institutional reports and across patient subgroups remains to be performed. Several tasks contain few questions, RSNA-RadioQA 79 and some knowledge tasks fewer after the radiology filter, so their individual estimates are less precise, whereas averages over questions are dominated by the IU/Open-i chest radiograph reports; the primary outcome, the task mean, weights every task equally. The questions on imaging orders, triage and follow-up were labeled by the agreement of two LLMs and were used only to augment the training data. The LLM comparator, Qwen3.8-27B, was evaluated zero-shot with a single prompt and was not fine-tuned on the same data, so the contributions of the decision format and of domain fine-tuning are not separated. Each model was trained once, with a single random seed. Public examination questions and Eurorad cases may have been part of the pretraining data; because Qwen3.8-27B, Kev-27B and RadKev-27B share the same backbone, such exposure is common to the principal comparisons. Latency was measured with one request at a time, and batched serving may increase the throughput of every system. All questions were posed on text.

### 4.3 Future work

The specialized model should be compared with radiologists in a reader study on institutional reports, with answer spaces taken from the institution’s own catalogs, and its error-detection and appropriateness decisions tested on reports and orders from other institutions. An LLM fine-tuned on the same data would separate the effect of the decision format from that of fine-tuning, and repeated training with different seeds would quantify the variability of the gains. Methods of constructing answer spaces should be compared by their inclusion rate and the accuracy and calibration they yield, routing should be evaluated on the information available before the read, and extending the approach to images would address the larger part of radiological practice.

## 5 Conclusions

Specializing an open, general-purpose decision model on public radiology data improved its accuracy on a radiology benchmark: the specialized 27B model was more accurate than its starting point, two hosted general-purpose decision models and its own backbone used as an LLM, with or without reasoning, at a fraction of the latency of reasoning, and the specialized 9B model did not differ detectably from a general-purpose model three times its size. The gains were concentrated in a few tasks and, for case diagnosis, reflected recognition of the answer options rather than reading of the case; the frequency of answers in the training data shaped the models’ preference for a residual option, and hedged findings, rare in the training data, remained a weakness on reports from another institution, where the 27B model lost accuracy. For categorical radiology decisions that must run within an institution, a specialized open decision model is a practical alternative to larger general-purpose models and to LLMs, provided that its training answers and answer spaces are curated for the intended use and that it is validated on local data.

## Data Availability

The code for data construction, teacher labeling, training, evaluation and statistical analysis, and the script that generates every number, table and figure in this manuscript from the job outputs, are available at https://github.com/udiram/RadKev. The RadKev-27B and RadKev-9B adapters and pointer heads are released on Hugging Face (https://huggingface.co/ramu9703/radkev-27b and https://huggingface.co/ramu9703/radkev-9b) under CC BY-NC-SA 4.0 for non-commercial research use, because several training sources carry that license; access requires acceptance of these terms. The Kev code on which they depend is distributed under Apache-2.0 [15].

https://github.com/udiram/RadKev

## Declarations

## Data availability

All data used in this study are publicly available. IU/Open-i [22] is distributed under CC BY-NC-ND 4.0, Eurorad [26, 27] under CC BY-NC-SA 4.0, MedMCQA [30] under Apache-2.0, MedQA [31] under CC BY 4.0, and MMLU [32], PubMedQA [33] and MedXpertQA [34] under MIT licenses; CT-RATE [23] (CC BY-NC-SA 4.0), CheXpert Plus [36] and ReXGradient-160K [37] are available from their providers under access agreements. The RadCases labels [6] are released under the MIT license with openly available case texts [29], ReXErr [24] is available from PhysioNet under the Open Data Commons Attribution License, the RSNA-RadioQA case descriptions are published in the supplement of the original article [8], and RadGraph-XL [21] is available from the Stanford Center for Artificial Intelligence in Medicine and Imaging under its research use agreement. The question records built from Eurorad, MedMCQA, MedQA, MMLU, PubMedQA and MedXpertQA are released in a Hugging Face dataset (https://huggingface.co/datasets/ramu9703/radkev-data), each under the license of its source. Because the license of IU/Open-i does not permit derivative works and the access-controlled sources may not be redistributed, records built from them, and those built from RSNA-RadioQA, are not released as text; instead, the record identifiers (report, case or patient), SHA-256 hashes of the record text, the ground-truth labels, the teacher labels and a script that rebuilds the records from the original datasets are provided. Per-question model outputs (probabilities for every answer option, without text) are released for all systems and analyses.

## Ethics statement

This study used only publicly available, de-identified datasets under their respective licenses and access agreements. No patients were recruited and no institutional patient data were used; review by an institutional review board was therefore not required.

## Author contributions

U.R. conceived the study, designed the methodology, wrote the software, built the datasets, trained and evaluated the models, performed the statistical analysis, prepared the figures and wrote the manuscript. R.Z. supervised the study and reviewed and edited the manuscript. Both authors approved the final manuscript.

## Use of AI tools

AI assistants (Claude, Codex) were used under the authors’ direction to write and run analysis code, to generate figures and to proofread and revise the final manuscript text. The authors reviewed all code, results and text and take full responsibility for the content.

## Acknowledgments

Training and evaluation were run on a workstation of the Zhang laboratory with four NVIDIA RTX A6000 GPUs. We thank the creators of IU/Open-i, Eurorad, CT-RATE, ReXErr, MIMIC-CXR, RadCases, RSNA-RadioQA, RadGraph-XL, CheXpert Plus, ReXGradient-160K, MedMCQA, MedQA, MMLU, PubMedQA and MedXpertQA for making their data available, the European Society of Radiology and the authors of the Eurorad teaching cases, and the developers of Kev, Qwen and MedGemma for releasing their models openly.

## Supplementary Information

## Supplementary Note S1. Example records

For each open-license source, the record shown is the one at the first quartile of state length among a fixed sample of 60 records per source, reproduced as the models received it. The correct option is shown in bold. For CT-RATE and the teacher-labeled questions, whose report text cannot be redistributed, the question formats are shown instead.

**IU/Open-i chest radiograph report** (development and test only; CC BY-NC-ND 4.0).

*State.* EXAM: Chest radiograph

INDICATION: SOB

COMPARISON: None available

FINDINGS: Flattening of the bilateral hemidiaphragms. Lungs are clear. Soft tissues and bony structures unremarkable. No pneumothorax or effusion.

IMPRESSION: Mild hyperexpansion. No acute process.

Q1. Is a lung nodule reported as present? *(yes/no) Answer:* no.

Q2. Does the radiologist report scoliosis in this study? *(yes/no) Answer:* no.

Q3. Is this a normal study with no abnormal findings? *(yes/no) Answer:* no.

**Eurorad case report** (case 14824, *Abdominal wall endometriosis: MRI findings and the “Gorgon sign”*, https://www.eurorad.org/case/14824; © European Society of Radiology, CC BY-NC-SA 4.0).

*State. Age:* 36

*Sex:* female

*Clinical history:* A 42-year-old female patient suffering from a painful “lump” at the anterior abdominal wall in the left lower quadrant. Physical examination revealed local tenderness, circumscribed hard-consistency nodule fixed at abdominal wall planes.

*Imaging findings:* Ultrasound (Fig.1) showed a sizeable (4×3×2.5 cm) hypoechoic mass with lobulated contour, which extended ventrally from the aponeurosis of the left rectus abdominis muscle into the subcutaneous fat. The latter structures showed pre-contrast T1-hyperintensity consistent with recent blood and a fluid-fluid level.

Additionally, several linear bands were seen radiating into the adjacent fat, configurating the characteristic “Gorgon sign”. After intravenous gadolinium (Fig.3) both the mass and radiating strands showed marked contrast enhancement.

Q1. Which diagnosis best explains the clinical history and imaging findings? *Options:* **opt_1: Scar endometriosis at the anterior abdominal wall**; opt_2: Abdominal wall (incisional) hernia; opt_3: Post-surgical seroma /haematoma; opt_4: Soft tissue malignancy e.g. sarcoma; opt_5: Bland (fibrotic) scar / keloid; opt_6: Rectus muscle desmoid; opt_7: Abdominal wall abscess; opt_8: Other benign soft tissue tumour e.g. lipoma; opt_9: Suture granuloma / gossypiboma. *Answer:* opt_1.
Q2. Which reading worklist should this study go to? *Options:* abdominal_imaging; neuroradiology; musculoskeletal_system; chest_imaging; uroradiology_genital_male_imaging: Kidneys, urinary tract, male genital organs; paediatric_radiology: Children; head_neck_imaging; cardiovascular: Heart and vessels; **genital_female_imaging: Female pelvis and reproductive organs**; breast_imaging: Breast; interventional_ radiology: Image-guided procedures. *Answer:* genital_female_imaging.

**MedMCQA examination question** (Apache-2.0).

*State. Subject:* Physiology

*Question:* Heart muscle, true are all except:

Q1. Which of the options is right? *Options:* **opt_1: Has multiple nuclei**; opt_2: Act as syncitium; opt_3: Has branching; opt_4: Has gap junctions. *Answer:* opt_1.

**MedQA examination question** (CC BY 4.0).

*State. Question:* A 49-year-old man comes to the physician because of a 1-week history of diarrhea and abdominal bloating. His stools are bulky, foul-smelling, and difficult to flush. Over the past 6 months, he has had recurrent dull epigastric pain that is triggered by meals and lasts for a few days. He drinks 6 to 8 beers daily. Abdominal examination shows mild epigastric tenderness with no rebound or guarding. A CT scan of the abdomen is shown. The structure indicated by the arrows is most likely lined by which of the following?

Q1. Which option correctly answers the question? *Options:* opt_1: Pyogenic membrane; opt_2: Columnar mucinous epithelium; **opt_3: Granulation tissue**; opt_4: Simple ductal epithelium. *Answer:* opt_3.

**MMLU examination question** (development and test only; MIT).

*State. Subject:* clinical knowledge

*Question:* Which of the following molecules does not form part of DNA?

Q1. Which of the options is right? *Options:* opt_1: Deoxyribose; **opt_2: Amino acid**; opt_3: Pyrimidine; opt_4: Purine. *Answer:* opt_2.

**PubMedQA research question** (development and test only; MIT).

*State. Research question:* Does screening or surveillance for primary hepatocellular carcinoma with ultrasonography improve the prognosis of patients?

*Abstract:* The purpose of this paper is to evaluate the efficacy of ultrasonographic screening for primary hepatocellular carcinoma. A total of 680 eligible cases were classified into three groups (surveillance, opportunistic, and symptomatic groups) according to their initial exposure. We used survival time, tumor morphology, and T staging as prognostic outcomes. The outcomes of screened/unscreened and sur veillance/nonsur veillance were compared with the use of the logistic regression model. The adjusted odds ratios for the screened group versus the unscreened group, with 1-, 2-, and 3-year survival time being used as outcomes, were 0.33 (95% confidence interval [CI], 0.21-0.52), 0.33 (95% CI, 0.21-0.53), and 0.37 (95% CI, 0.23-0.61), respectively. The adjusted odds ratios for surveillance versus nonsurveillance were 0.58 (95% CI, 0.35-0.97), 0.45 (95% CI, 0.27-0.74), and 0.44 (95% CI, 0.26-0.73). The odds ratios were even smaller when tumor morphology or T stage was taken as the main outcome. All these results were statistically significant. There were significant gradient relationships between prognostic outcomes and extent of screening history.

Q1. What do the study’s results say about the question? *Options:* **yes: Yes, the evidence supports it**; no: No, the evidence does not support it; maybe: Maybe: the evidence is mixed or inconclusive. *Answer:* yes.

**MedXpertQA examination question** (development and test only; MIT).

*State. Question:* A 54-year-old woman with a history of diabetes and peripheral vascular disease presents with swelling and aching pain in her calves that worsens after prolonged sitting. On examination, her posterior tibialis and dorsalis pedis pulses are graded 2/4 bilaterally. Both legs are symmetrically enlarged and non-pitting, with a negative Homan’s sign on both sides. General tenderness is noted from the ankles to the knees without localized areas of pain. To directly address her condition, what is the most appropriate next step in treatment?

Q1. Choose the correct answer. *Options:* opt_1: Venous pump activation exercises; opt_2: Pedal pump technique; opt_3: Thoracic inlet release technique; opt_4: Pelvic diaphragm release technique; opt_5: Lower extremity muscle energy technique; opt_6: Effleurage of the bilateral legs; opt_7: Lymphatic drainage massage; opt_8: Myofascial release of the gastrocnemius; opt_9: Compression stockings application; **opt_10: Indirect thoracoabdominal diaphragm release**. *Answer:* opt_10.

**CT-RATE chest CT report** (CC BY-NC-SA 4.0; access-controlled, report text not reproduced).

*State.* A CT-RATE chest CT report with clinical information, technique, findings and impression.

Q1. Does this report describe a pleural effusion? *(yes/no; three such questions per report)*

Q2. Is this a normal study with no abnormal findings? *(yes/no)*

Q3. Which of these findings is described in the report? *(one finding present and three absent)*

Answers follow the dataset’s classifier labels.

**Teacher-labeled questions** (states from CheXpert Plus, ReXGradient-160K and CT-RATE reports and from clinical indications, including Eurorad case presentations).

*Report question.* State: a radiology report. What follow-up does this report call for? *Options:* No further imaging recommended; Follow-up imaging within 3 months; Follow-up imaging in more than 3 months; Further evaluation with a different imaging test or procedure; Clinical correlation or non-imaging workup only.

*Order question.* State: the clinical indication only. Which imaging study is most appropriate first for this clinical question? *Options:* Chest radiograph; CT head without contrast; CT pulmonary angiography (PE protocol); CT chest; CT abdomen and pelvis with IV contrast; Ultrasound abdomen (incl. RUQ); Ultrasound pelvis / transvaginal; MRI brain; MRI spine; Radiograph of the painful bone or joint; MRI of a joint or limb; Doppler ultrasound of vessels; Mammography and/or breast ultrasound; No imaging indicated.

The label is the option that both teachers ranked first; the training target is the mean of the two teachers’ distributions.

**Question formats of ReXErr, RadCases and RSNA-RadioQA. *ReXErr*** (state: a chest radiograph report, either the original or its error-injected version): “Does this report contain an error?”, or one of three alternative wordings, answered yes or no. *RadCases* (state: a one-sentence patient summary): “Which ACR Appropriateness Criteria panel covers the imaging work-up of this patient?”, with the 12 options formed by the panels and “None: no ACR Appropriateness Criteria topic applies”, and “Which ACR Appropriateness Criteria topic best matches this patient’s presentation?”, with the 225 topics as options. *RSNA-RadioQA* (state: the case description): “What is the most likely diagnosis?”, with four diagnoses as options. The case descriptions of RSNA-RadioQA are not reproduced.

## Supplementary Note S2. Record construction

### IU/Open-i

Medical Subject Headings assigned by the indexers were mapped to 14 findings. A finding was considered present if one of its terms had been assigned and absent otherwise, and a study was considered normal if it had been indexed as normal. Reports with fewer than 20 characters of findings and impression, and reports without indexing, were excluded. Each report received two presence questions, chosen so that one finding was present and one absent where possible, one normal-study question and, for half of the reports with at least one finding, a question on which of four findings (one present, three absent) was described. The license (CC BY-NC-ND) does not permit derivative works, so the collection was used only for development (25% of reports, by a hash of the report identifier) and testing (75%).

### CT-RATE

The collection comprises 25,692 chest CT studies from 21,304 patients [23]; its labels were produced by a RadBERT classifier fine-tuned on manually annotated reports. One report per study received three presence questions and the normal-study and which-finding questions; a study was normal if none of the 18 abnormalities was labeled. The official training set was divided by patient into training and development records (8% development), the official validation set served as the test set, and training used a random sample of 8,000 reports.

### Eurorad

The state contained age, sex, clinical history and imaging findings, from which sentences mentioning pathology, biopsy, histology, surgery, treatment, follow-up, confirmation or diagnosis were removed by keyword matching. Cases whose final diagnosis did not appear among the differential diagnoses, that had fewer than two differential diagnoses or whose remaining findings were shorter than 40 characters were excluded, as were development and test cases whose remaining text contained the diagnosis. Half of the cases received the subspecialty question, whose correct answer was the Eurorad section of the case (the first listed when several were given) and whose options were shown with a one-line description of the section in 60% of cases. Because the state contained the imaging findings, this question classifies an interpreted case; it does not reproduce the routing of a study before interpretation, which relies on the order, the indication and the examination type. Age and sex were taken from the case metadata and occasionally differ from those stated in the history, as in the example of Supplementary Note S1. Cases were split 80%, 10% and 10% by a hash of the case identifier.

### RSNA-RadioQA

The case descriptions and reference answers were taken from Appendix S1 of the original study [8], which omits question 44 of the 80, and the four answer options of each question from the public RadSaFE-200 release (RadioRAG subset), which contains the options of the RaR study [28]. The state was the case description and the question its final sentence, which asks for the most likely diagnosis; options were presented under neutral identifiers in a fixed random order. The correct answer was the option matching the reference answer of the appendix, which every question contained; for 2 questions, the answer index of the option release designates a different option, and the reference answer was used.

### RadCases

The RadCases release (Hugging Face michaelsyao/RadCases, revision 93c99559) labels each patient summary, a one-sentence description of the presentation, with the ACR Appropriateness Criteria panel and topic that apply to it; the labels were assigned by two senior medical students under the supervision of an attending radiologist, with disagreements resolved by radiologists [6]. The release contains only SHA-512 hashes of the summaries. Their text was rebuilt from the two openly available subsets, summaries written by GPT-3.5 for the original study and the first sentences of Medbullets USMLE Step 2 and 3 questions [29], with the sentence splitter of the original study, and a case was retained only if the hash of its rebuilt text matched a label (468 of 471). Summaries whose text occurs in any other record of this study (37) were excluded, and the remaining cases were split by a hash of the summary into training (60%), development (10%) and test (30%) cases. Each case received the panel question unless it was labeled with more than one panel (12 cases) and the topic question if it was labeled with exactly one known topic. The panel question offered the 11 panels and the option “None: no ACR Appropriateness Criteria topic applies”, the topic question all 225 topics of the criteria version used by the original study, both under neutral, shuffled identifiers. The state was the summary alone.

### ReXErr

ReXErr-v1 [24] pairs MIMIC-CXR reports [25] with versions into which GPT-4o inserted clinically meaningful errors, sampled across error types that occur in reports written by radiologists and by generative models. At the report level, each study was represented by one record whose state was either the original or the error-injected report, chosen by a hash of the study identifier so that about half of the records contain an error, and whose single question asked whether the report contains an error; the ground truth is therefore known by construction. Studies whose two versions were identical or empty were excluded. The official splits were used, with 6,000 training and 500 development reports sampled at random and all 2,708 test reports retained (1,376 with an error).

### Examination questions

MedMCQA: all radiology questions were retained; other subjects were sampled at approximately 20% and capped at 2,500 training questions per subject; questions with empty or duplicate options were removed; because the official test answers are not public, the official validation set served as the test set, and 5% of the official training set was held out for development. MedQA: the official test set was retained and 10% of the official training set was held out for development. MMLU: the anatomy, clinical knowledge, college biology, college medicine, medical genetics and professional medicine subjects, with the official validation and test sets used for development and testing. PubMedQA: the expert-annotated subset, divided equally between development and testing. MedXpertQA: the text subset, 20% for development and 80% for testing. For the present models, every split of these collections was restricted to radiology: all MedM-CQA radiology questions were retained, and a question from any other subject or collection was retained if its stem, state or options contained a term from a fixed list of imaging modalities and radiological terms (for example radiograph, X-ray, computed tomography, magnetic resonance, ultrasound, mammography, scintigraphy, angiography, fluoroscopy, contrast agent, imaging, radiology and radiopaque), with acronyms such as CT, MRI, PET and SPECT matched only in capitals. The list was fixed before any result of the present models was read.

### Teacher-labeled questions

Up to 1,500 reports and 1,500 indications were drawn per source, including Eurorad case presentations for indications. Five kinds concern a report (critical finding, urgency, follow-up, interval change and incidental finding), one the status of a finding in a chest radiograph report, and four an imaging order (first examination, use of contrast, priority and appropriateness of an ordered examination); order questions were posed on the clinical indication alone. Each teacher was instructed to answer as an attending radiologist, with reasoning disabled and the options as a lettered list. Because MedGemma’s chat template does not suppress its reasoning segment, a one-sentence segment (“I will answer with the letter only.”) was pre-filled so that the next token is the answer. In the run whose labels were used for training, agreement ranged from 28.1% (use of contrast) to 84.7% (finding status). The labels used for training come from a first run in which MedGemma was scored without the pre-filled segment (22,557 questions retained, 68.4%). All candidates were later rescored by MedGemma with the pre-filled segment and the labels regenerated, with the answers of Qwen3.8-27B unchanged. Of the 19,945 training questions retained in the first run, 18,559 (93.1%) were also retained in the regenerated run, and no question retained in both runs received a different label. RadKev-27B and RadKev-9B were trained with the labels of the first run.

**Table S1.**
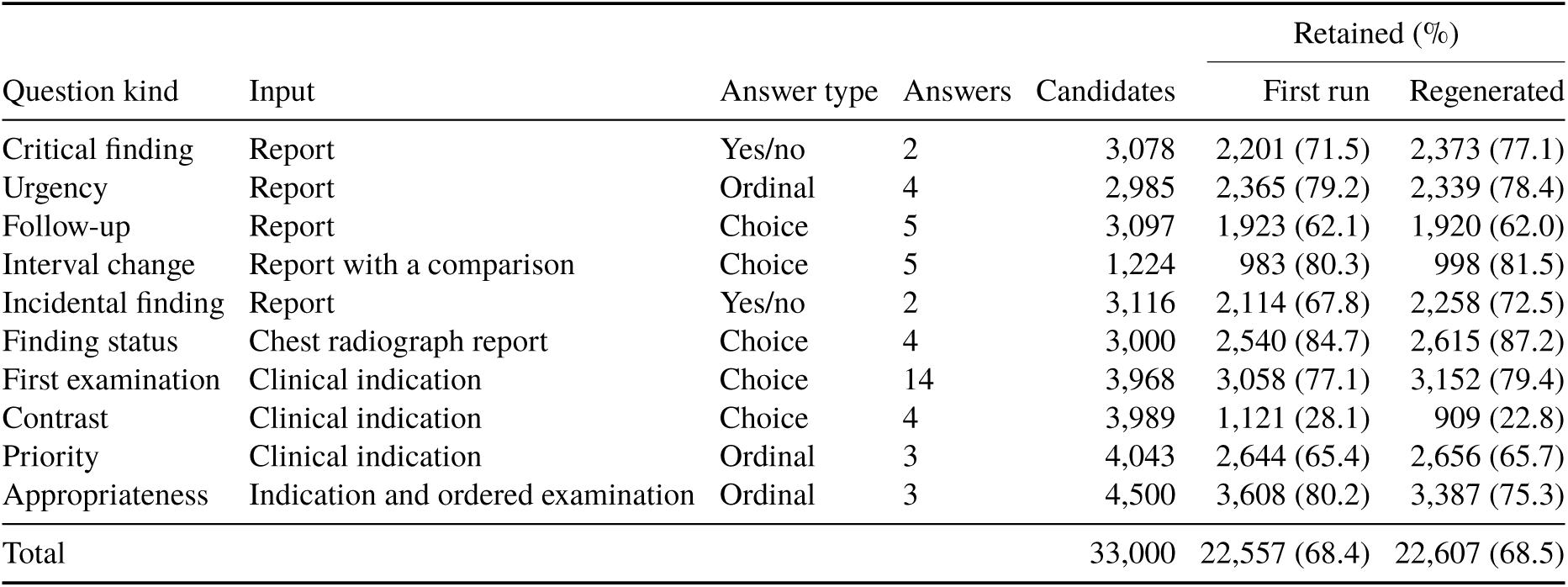
Teacher-labeled question kinds. Candidates were answered by MedGemma-27B-text and Qwen3.8-27B; a question was retained when both ranked the same answer first. Order questions saw only the clinical indication. First run, labels used for training, model selection and the main evaluation; regenerated, labels after rescoring MedGemma with the pre-filled reasoning segment.

## Supplementary Note S3. Training details

### Augmentation

Each multiple-choice question with a single correct answer received at most one modification: the correct option was replaced by a “none of the above” option, which then became the correct answer (probability 0.10; questions with more than two options); a “none of the above” option was added as an incorrect alternative (probability 0.12); or an irrelevant distractor was added (probability 0.15). With probability 0.25, a record additionally contributed a minimal pair in which the same question was posed with a “none of the above” option, once with and once without its correct option. With the 1,000 replayed Kev records, training comprised 36,954 records and 44,068 examples for RadKev-27B (44,070 for RadKev-9B), including the minimal pairs.

### Optimization

AdamW with weight decay 0.01; one-cycle schedule with 10% warm-up and cosine decay; gradient-norm clipping at 1.0; 8 records per optimizer step, from micro-batches of 2 records across data-parallel processes and accumulation steps; gradient checkpointing; states limited to 1,536 tokens, question branches to 2,176 tokens and packed records to 3,200 tokens, limits that 155 training records exceeded and that excluded them from training; random seed 0. The temperature was selected from 121 logarithmically spaced values between 0.25 and 4.

### Models

Kev-27B, Hugging Face revision 01b8199, on Qwen3.8-27B revision 1d4bf0f; Kev-9B, revision 2629c06, on Qwen3.5-9B-Base revision 68c46c4. The pointer head projects the hidden states of each option’s closing token and of the question’s decision token into a 256-dimensional space, and the logit of an option is the scaled dot product of the two projections.

### Hardware and software

Peak memory, recorded on the first GPU of the first process, was 33.5 GiB for RadKev-27B and 42.5 GiB for RadKev-9B. For Kev-9B, splitting the model by layers across two GPUs reproduced single-GPU probabilities exactly on 74 development questions. Software: Kev at commit f2bb629 with two local patches, released with the code, that split a model across a pair of GPUs and add data-parallel training of the adapter (summation of gradients across processes, saving by the first process and a periodic check that all processes hold identical weights, which passed at each of the 10 checks of both runs), PyTorch 2.8.0 with CUDA 12.8, Transformers 5.17.0, PEFT 0.21.0, flash-linear-attention 0.5.2 and causal-conv1d 1.7.0.

## Supplementary Note S4. Evaluation details

### Decision models

Each record was submitted once, with all of its questions, in bfloat16; each model used its own temperature (the released temperature for the general-purpose models and the fitted temperature for RadKev). The answer to a question is the option with the highest probability, and that probability is its confidence. Every model accepted every test question.

### Language models

The prompt was that used for teacher labeling: the state, the question and its options as a lettered list, followed by an instruction to answer with one letter. With reasoning enabled, Qwen3.8-27B was run in its thinking mode using vLLM 0.19 in bfloat16, the model’s default sampling settings, seed 0 and at most 4,096 reasoning tokens. After the reasoning, “Answer:” was appended, and the answer distribution was read from the next-token probabilities of the option letters. The reasoning sample comprised 1,675 benchmark questions, selected in a fixed hash order that does not depend on any model output: every Eurorad and MedMCQA radiology question, up to 200 questions from each IU task, every RSNA-RadioQA and RadCases panel question, 200 ReXErr questions and, from each other examination source, the radiology questions among the first 60 questions in that order, which left between 1 (MedMCQA, other subjects) and 20 (MMLU) questions per task. For the task average of the reasoning sample, the questions of the six examination tasks were therefore pooled into one task, giving nine tasks; bootstrap resampling was stratified by source, as in the main evaluation. Averaged over the 14 tasks without pooling, the difference between RadKev-27B and Qwen3.8-27B with reasoning was 0.8 points (−1.6 to 3.1).

### Outcomes

The expected calibration error was computed over ten equal-width bins of confidence. A confident error is a question answered incorrectly with a confidence of at least 0.9. Coverage at a 5% error budget is the largest proportion of questions that can be accepted in descending order of confidence while the error among the accepted questions does not exceed 5%.

## Supplementary Note S5. Answer-space analysis

For each question, the state, the instructions and the human-labeled correct answer were held fixed, and only the other options were varied. The questions were 30 Eurorad diagnosis questions of the test split and 30 RSNA-RadioQA questions, the first in a fixed hash order of their identifiers. Pool distractors were drawn from all distinct Eurorad diagnosis options of the training, development and test splits, and for RSNA-RadioQA additionally from its own options, either at random or in order of decreasing similarity to the correct answer, measured as the cosine between PubMedBERT sentence embeddings [54]. LLM distractors were generated by Qwen3.8-27B, which received the state, the question and the correct answer and was asked, with reasoning disabled and greedy decoding, for 15 plausible but incorrect answers; the correct answer was never taken from the LLM. A candidate distractor was discarded if its normalized text contained, or was contained in, that of the correct answer or of an already selected distractor, or if its embedding cosine to either was at least 0.95. Each question was posed with its original options; with the correct answer and random or most similar pool distractors, for 2, 4, 16, 64 and 255 options in total; with the correct answer and LLM distractors, for 2, 4, 8 and 16 options; and with the original options supplemented with up to 8 LLM distractors. An answer space was omitted for a question when too few distractors remained after filtering. Each answer space was posed as a separate question on the same state. RadKev-27B, Kev-27B, RadKev-9B and Kev-9B scored every option directly. The LLM received the options as a numbered list and generated the number of its answer by greedy decoding, with reasoning disabled; an output that was not a number in range was counted as incorrect, and its rate is reported. Up to 16 options, the LLM was additionally scored by option-letter probabilities, as in the main evaluation. Accuracy was estimated with 95% intervals from a bootstrap over questions. Latency as a function of the number of options was measured for every system with one question per request on 15 Eurorad cases with 2, 16, 64 and 255 options, one request at a time, with the first 5 requests excluded; the LLM was timed with numbered generation at every size and with option-letter scoring at up to 16 options. In the main latency measurement, the first 5 requests of each mode (the first request for the LLM with reasoning) were excluded, and the per-question latency of a decision model was its latency per record divided by the number of questions.

## Supplementary Note S6. External test set

### RadGraph-XL

The Stanford portion of RadGraph-XL comprises 2,000 reports, 500 each of chest radiographs, chest CT, abdominal and pelvic CT and brain MRI, in which board-certified radiologists annotated every entity and its status [21]. Reports sharing at least half of their word 8-grams with a CheXpert Plus report or with any record of this study were excluded (495 reports, all of them chest radiograph reports, which derive from the same institution as CheXpert Plus), so that the test consists almost entirely of CT and MRI reports. For each remaining report, one observation annotated as definitely present, one as definitely absent and one as uncertain were drawn at random where available, excluding spans that consist only of a generic qualifier such as “normal” or “stable”, and each was posed in the format of the finding-status questions used in training, with the annotated span as the finding and the options reported as present, explicitly reported as absent, possible or equivocal, and not mentioned in the report; the last was never correct. The state was the full report text.

### Analysis

Accuracy and paired differences were estimated as in the main evaluation, with 2,000 bootstrap resamples of records stratified by modality, on all questions, on the questions about definite findings (with the uncertain option removed from every answer) and on the hedged findings. Supplementary Table S12 gives the accuracy of every system in each of these sets. The hosted decision models received each report once with all of its questions, as in the main evaluation, and neither API declined a question.

## Supplementary Tables

**Table S2.**
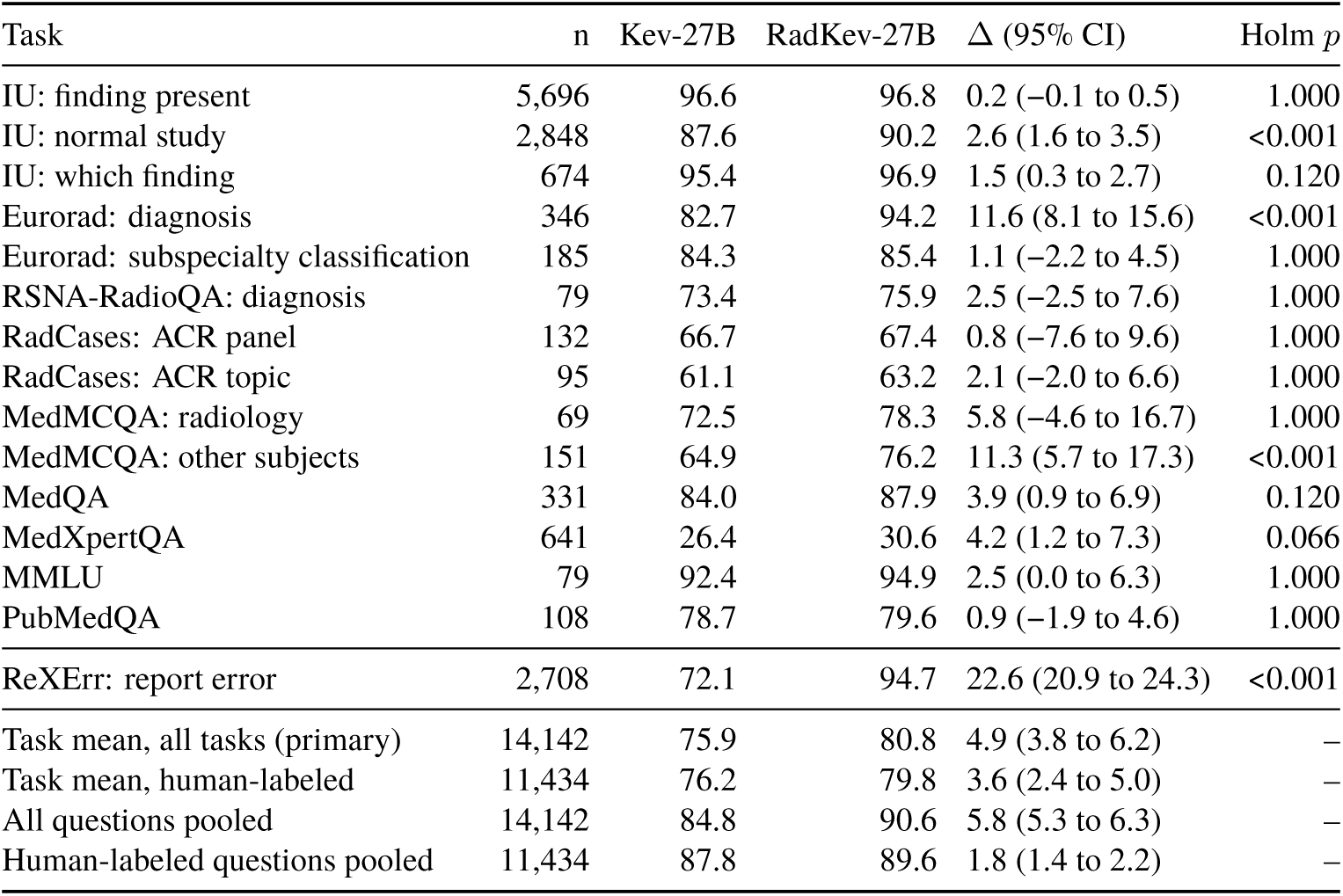
Primary comparison by task: accuracy (%) of Kev-27B and RadKev-27B and their difference in percentage points (95% confidence interval, paired cluster bootstrap with 2,000 resamples). Holm *p*: adjusted across the fifteen benchmark tasks. Top: tasks with human-labeled ground truth; middle: ReXErr, with ground truth known by construction; bottom: averages (the first row is the prespecified primary outcome).

**Table S3.**
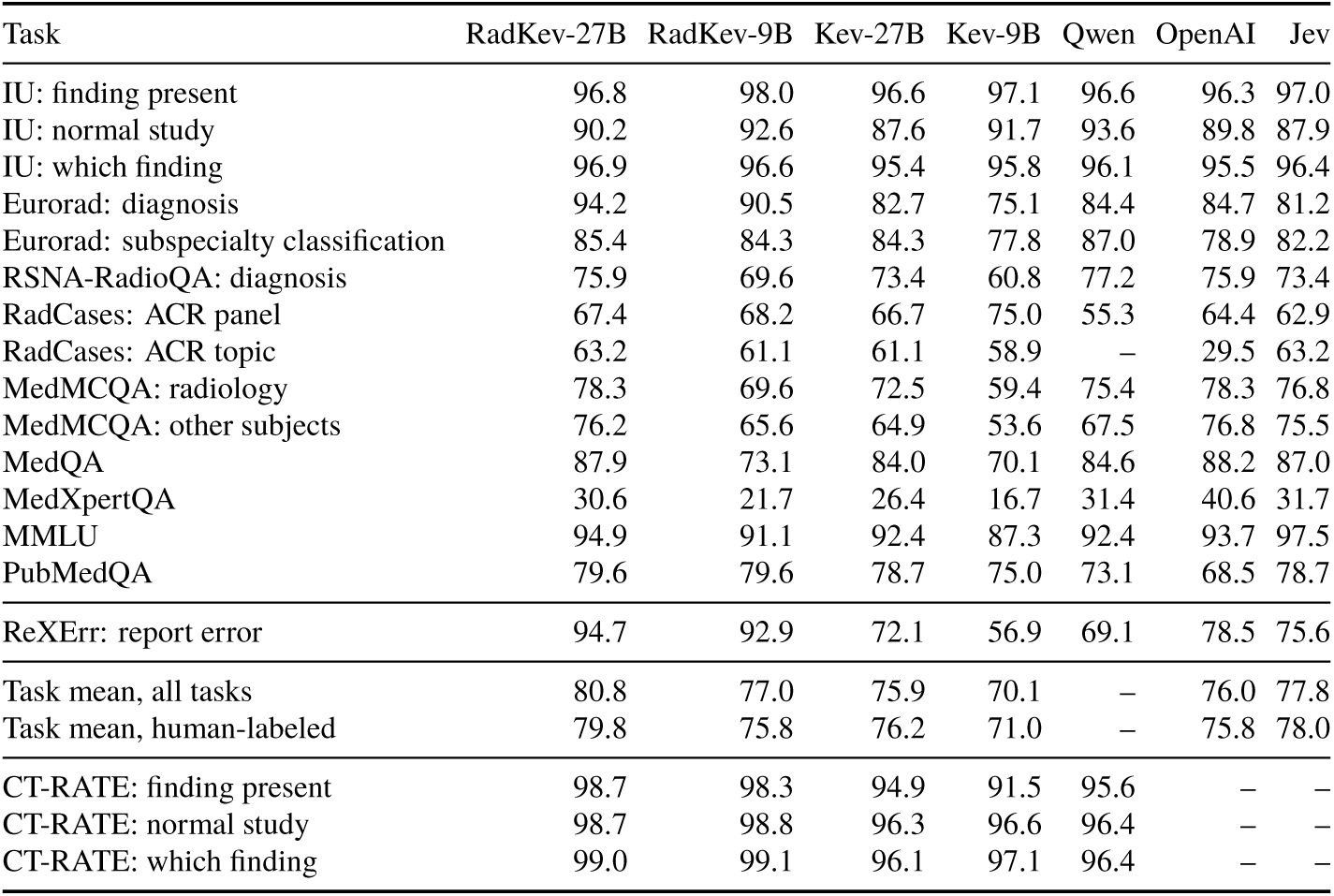
Accuracy (%) of every system on every benchmark task and, at the bottom, agreement with the CT-RATE classifier labels, which are not part of the benchmark. Qwen, Qwen3.8-27B, which was not scored on the RadCases topic question (225 options); a dash marks a value not computed.

**Table S4.**
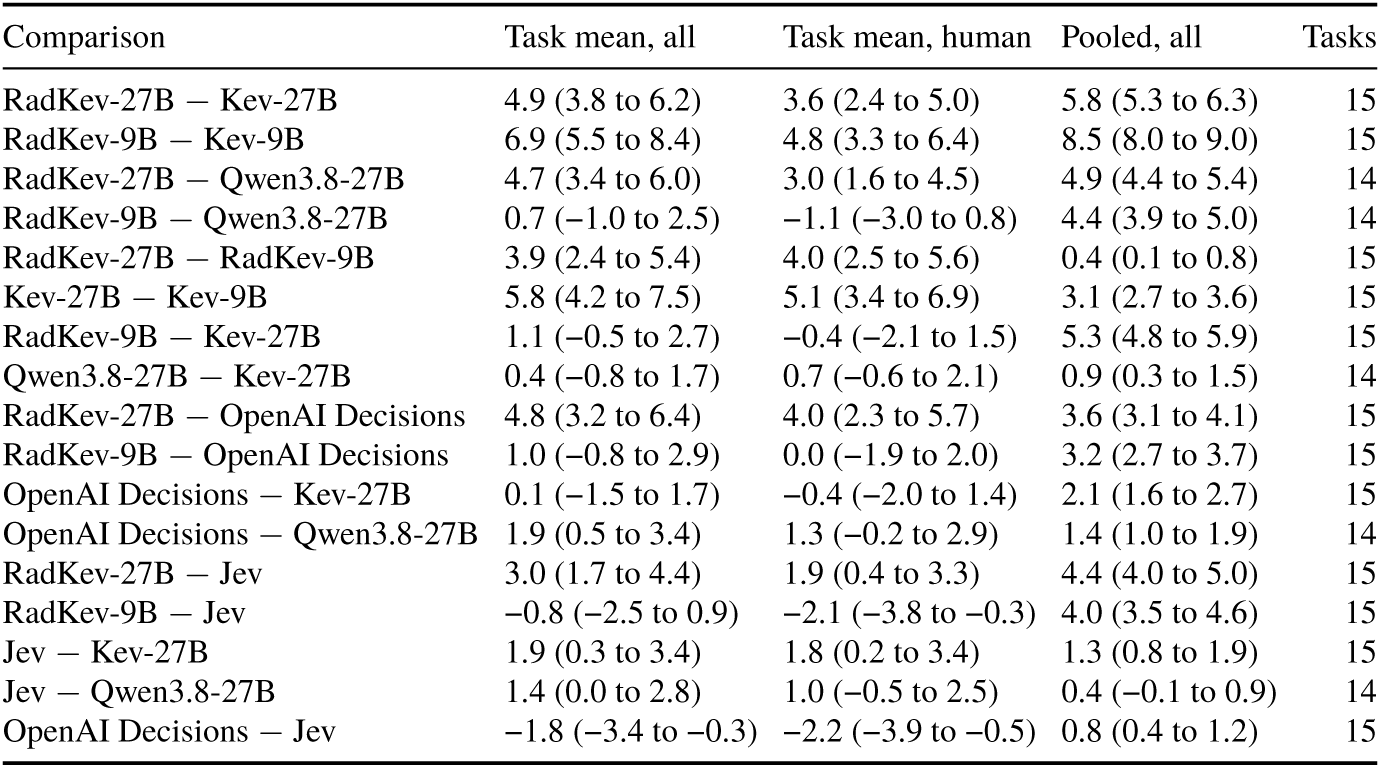
Paired comparisons in percentage points (95% confidence interval): task-averaged accuracy over all benchmark tasks and over the tasks with human-labeled ground truth, and accuracy pooled over all questions. Comparisons with an LLM use the questions with at most 16 options. Tasks: number of tasks compared.

**Table S5.**
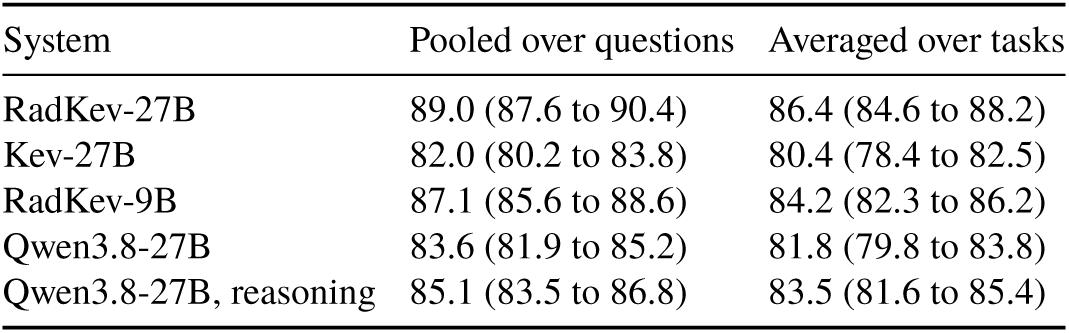
Accuracy (%, 95% confidence interval) on the reasoning sample of 1,675 benchmark questions, pooled over questions and averaged over tasks, with the questions of the six examination tasks pooled into one task, and with reasoning disabled and enabled for Qwen3.8-27B.

**Table S6.**
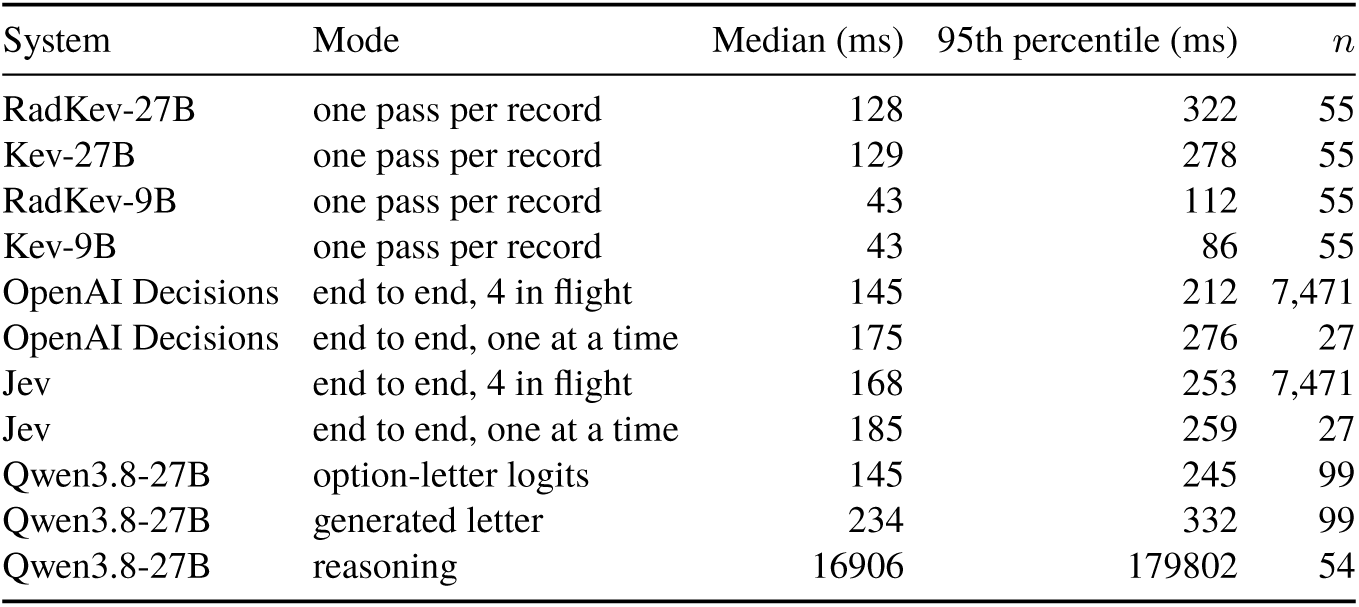
Latency per question on 60 benchmark records (one request at a time, two RTX A6000 GPUs, Hugging Face Transformers, bfloat16): median and 95th percentile. *n*: timed records for the decision models, including the OpenAI Decisions API and Jev, whose latency per question is the latency of the record divided by its number of questions, and timed questions for Qwen3.8-27B.

**Table S7.**
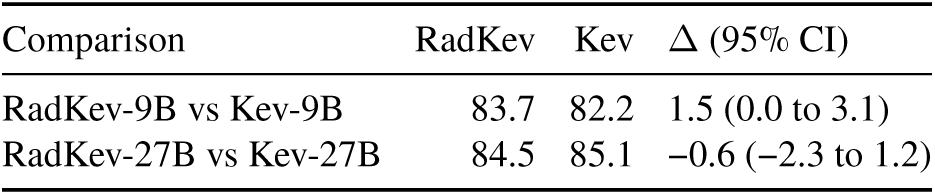
Kev’s out-of-domain transfer suite (656 questions): accuracy (%) of each specialized model and of the released model it started from, and their difference in percentage points (95% confidence interval).

**Table S8.**
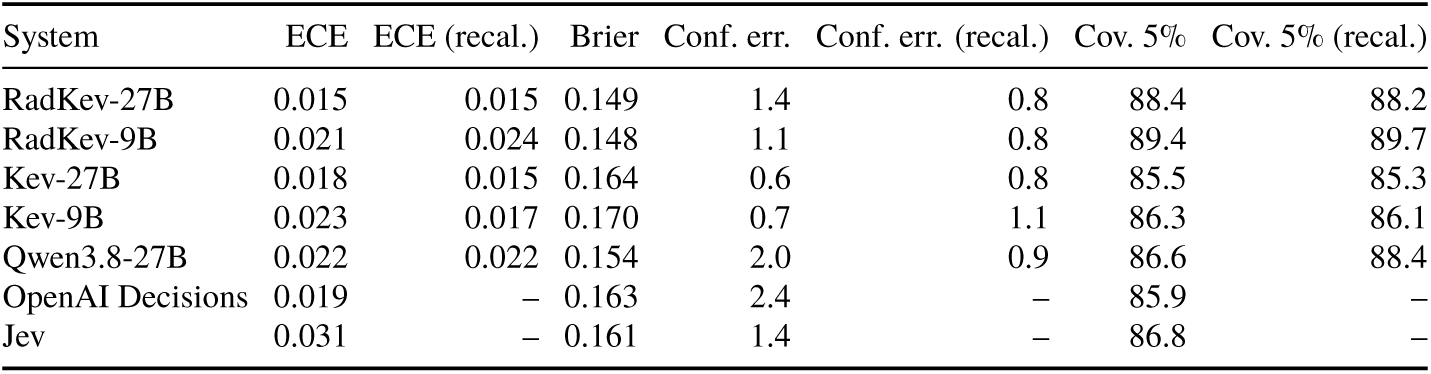
Calibration and selective prediction on the human-labeled questions: expected calibration error (ECE) over ten equal-width bins, Brier score, confident errors (% of questions answered incorrectly with a confidence of at least 0.9) and coverage at a 5% error budget (%), as scored and after two-fold cross-fitted temperature recalibration on the test split (recal.). The OpenAI Decisions API and Jev return probabilities rounded to two decimals, so they are reported as scored only.

**Table S9.**
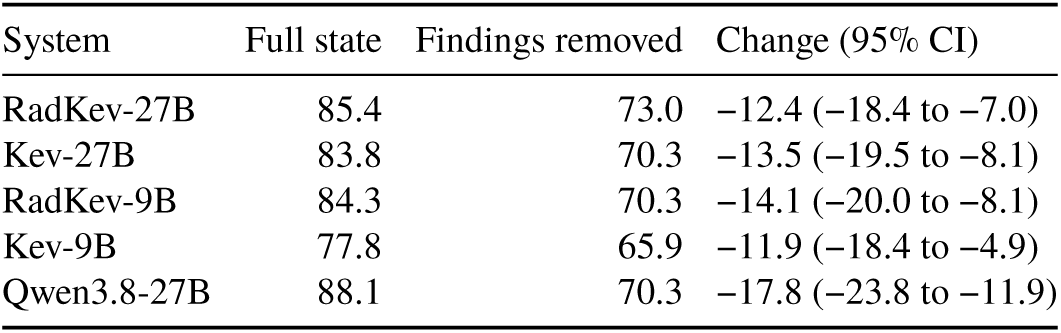
Eurorad subspecialty classification with the full state and with the imaging findings removed (age, sex and clinical history retained), as before the read: accuracy (%) and change in percentage points (95% confidence interval), 185 questions. Accuracies with the full state were recomputed in this analysis, with each question in its own request; they differ from the main evaluation by one question for Kev-27B and two for Qwen3.8-27B.

**Table S10.**
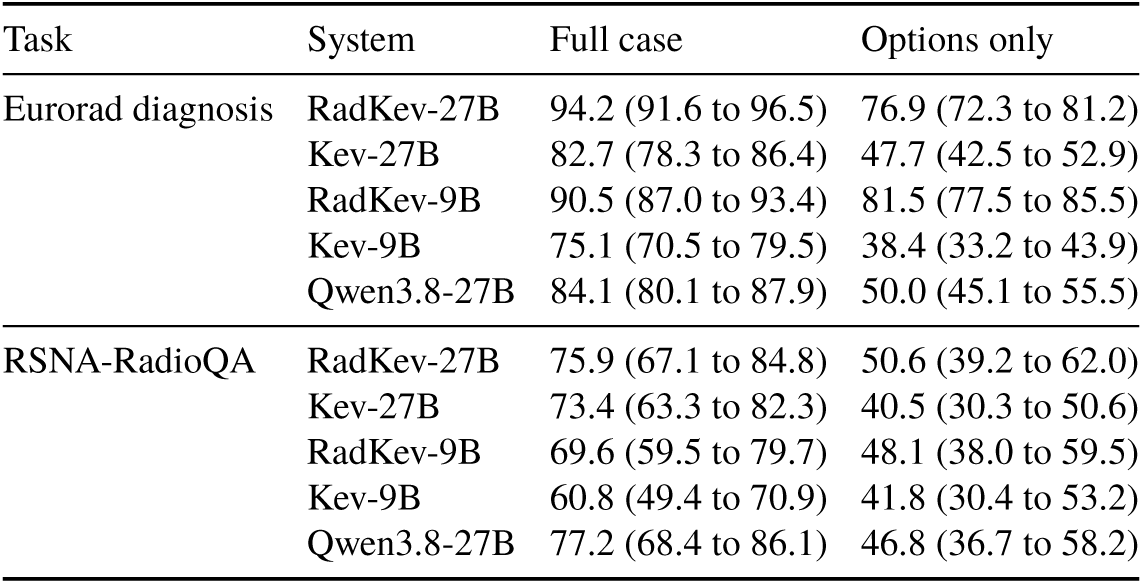
Options-only control: accuracy (%, 95% confidence interval) with the full case and with only the question and its options shown. Accuracies with the full case were recomputed in this analysis; for Qwen3.8-27B they differ from the main evaluation by less than 0.5 percentage points because of batch padding.

**Table S11.**
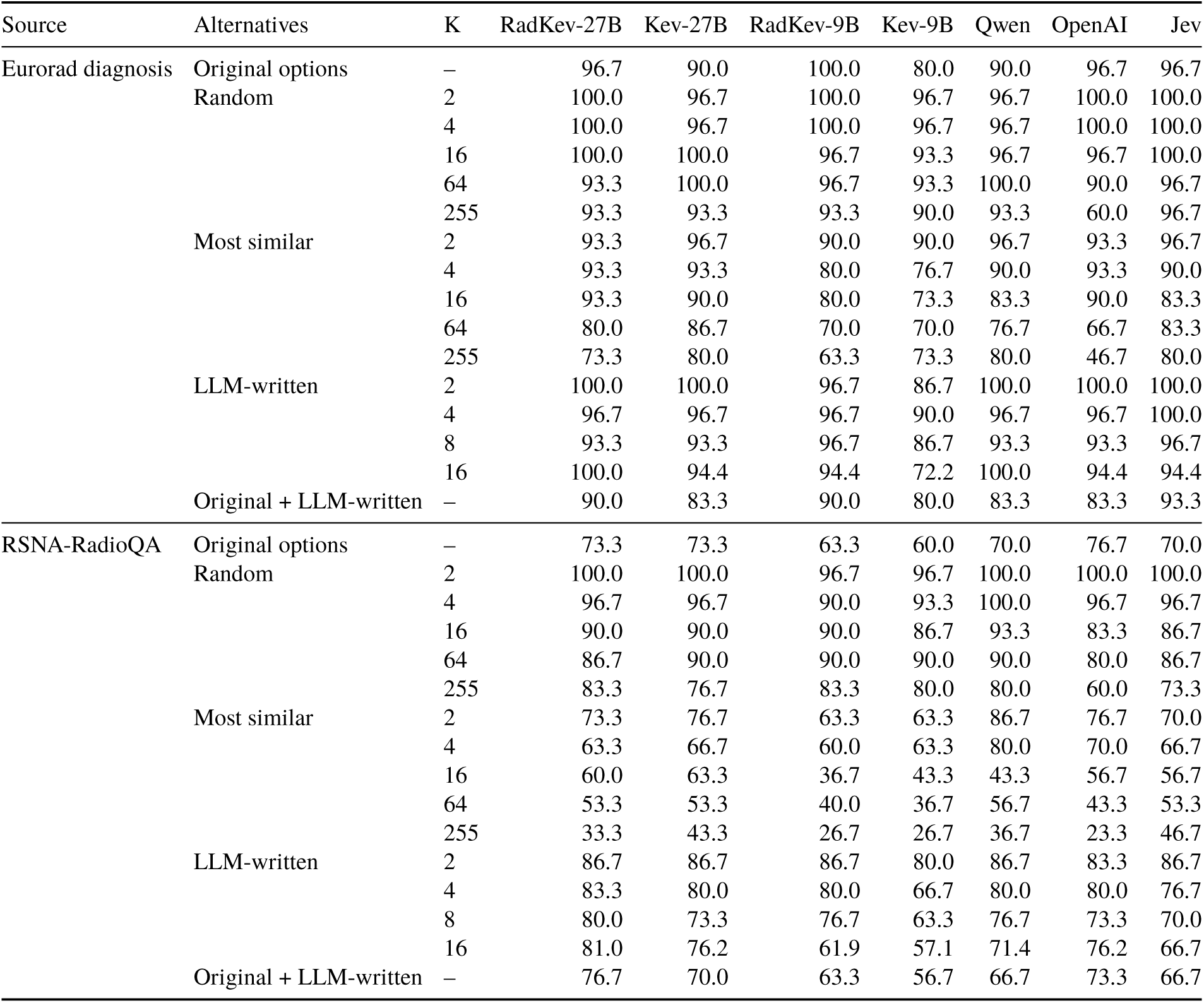
Accuracy (%) under each answer space on 30 Eurorad diagnosis and 30 RSNA-RadioQA questions; K, number of options offered. Random and most similar: alternatives drawn from the pool of diagnoses at random or in order of similarity to the correct answer. LLM-written: alternatives written by Qwen3.8-27B; with 16 options, 18 Eurorad and 21 RSNA-RadioQA questions retained enough alternatives after filtering. Original + LLM-written: the original options supplemented with up to 8 LLM-written alternatives. Qwen3.8-27B generated the number of the chosen option.

**Table S12.**
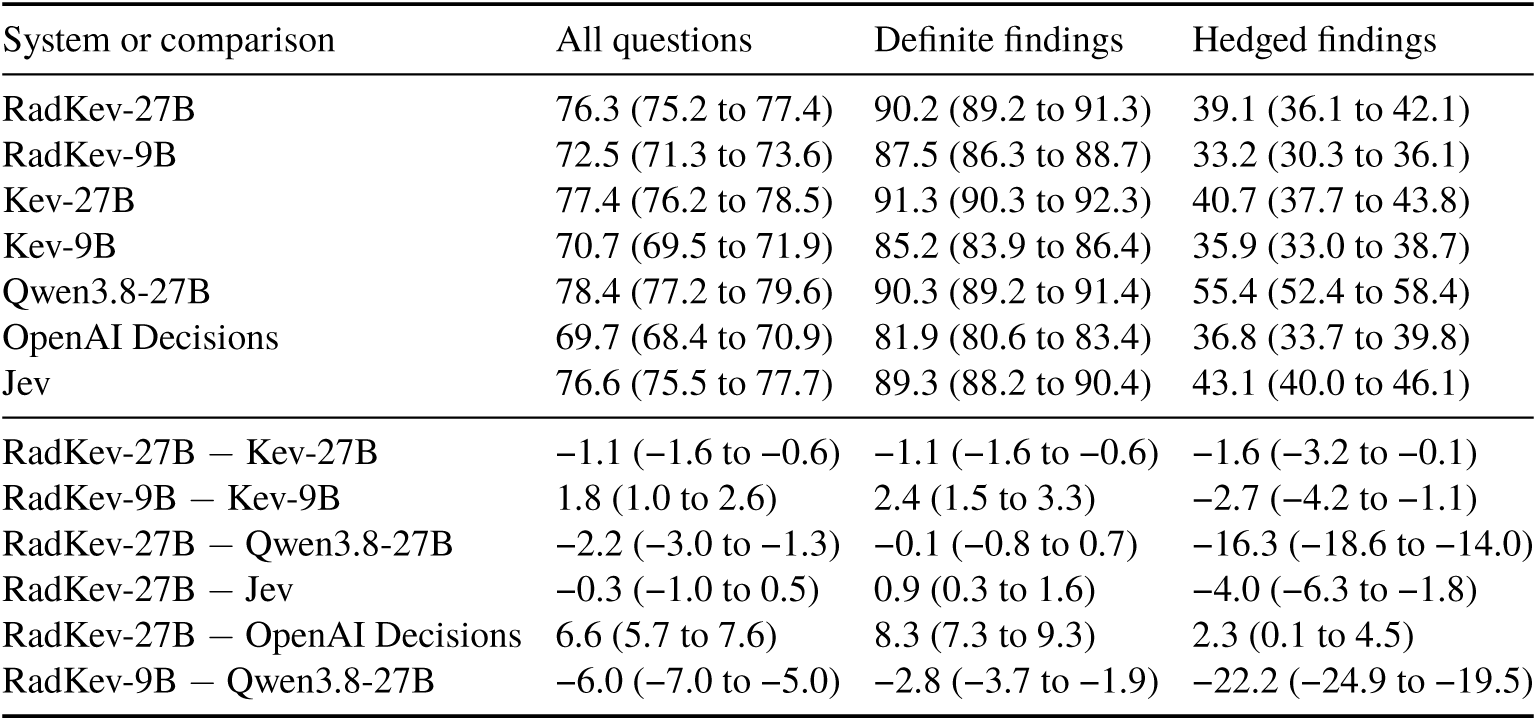
External test set (RadGraph-XL): accuracy (%, 95% confidence interval) and paired differences in percentage points on all 4,037 status questions, on the 2,991 questions about definite findings (present or absent; uncertain option removed from every answer) and on the 1,046 hedged findings (annotated as uncertain).

## References

[1] Yonatan Geifman and Ran El-Yaniv. Selective classification for deep neural networks. In Advances in Neural Information Processing Systems 30, pages 4878–4887, 2017. arXiv:1705.08500.

[2] Matthias A. Fink, Arved Bischoff, Christoph A. Fink, Martin Moll, Jonas Kroschke, Luca Dulz, Claus Peter Heußel, Hans-Ulrich Kauczor, and Tim F. Weber. Potential of ChatGPT and GPT-4 for data mining of free-text CT reports on lung cancer. Radiology, 308(3):e231362, 2023. doi: 10.1148/radiol.231362.

[3] Pritam Mukherjee, Benjamin Hou, Ricardo B. Lanfredi, and Ronald M. Summers. Feasibility of using the privacy-preserving large language model Vicuna for labeling radiology reports. Radiology, 309(1): e231147, 2023. doi: 10.1148/radiol.231147.

[4] Roman Johannes Gertz, Thomas Dratsch, Alexander Christian Bunck, Simon Lennartz, Andra-Iza Iuga, Martin Gunnar Hellmich, Thorsten Persigehl, Lenhard Pennig, Carsten Herbert Gietzen, Philipp Fervers, David Maintz, Robert Hahnfeldt, and Jonathan Kottlors. Potential of GPT-4 for detecting errors in radiology reports: Implications for reporting accuracy. Radiology, 311(1):e232714, 2024. doi: 10.1148/radiol.232714.

[5] Alexander Rau, Stephan Rau, Daniela Zoeller, Anna Fink, Hien Tran, Caroline Wilpert, Johanna Nattenmueller, Jakob Neubauer, Fabian Bamberg, Marco Reisert, and Maximilian F. Russe. A context-based chatbot surpasses trained radiologists and generic ChatGPT in following the ACR appropriateness guidelines. Radiology, 308(1):e230970, 2023. doi: 10.1148/radiol.230970.

[6] Michael S. Yao, Allison Chae, Piya Saraiya, Charles E. Kahn, Walter R. Witschey, James C. Gee, Hersh Sagreiya, and Osbert Bastani. Evaluating acute image ordering for real-world patient cases via language model alignment with radiological guidelines. Communications Medicine, 5:332, 2025. doi: 10.1038/s43856-025-01061-9.

[7] Daiju Ueda, Yasuhito Mitsuyama, Hirotaka Takita, Daisuke Horiuchi, Shannon L. Walston, Hiroyuki Tatekawa, and Yukio Miki. ChatGPT’s diagnostic performance from patient history and imaging findings on the Diagnosis Please quizzes. Radiology, 308(1):e231040, 2023. doi: 10.1148/radiol.231040.

[8] Soroosh Tayebi Arasteh, Mahshad Lotfinia, Keno Bressem, Robert Siepmann, Lisa Adams, Dyke Ferber, Christiane Kuhl, Jakob Nikolas Kather, Sven Nebelung, and Daniel Truhn. RadioRAG: Online retrieval-augmented generation for radiology question answering. Radiology: Artificial Intelligence, 7 (4):e240476, 2025. doi: 10.1148/ryai.240476.

[9] Rajesh Bhayana, Satheesh Krishna, and Robert R. Bleakney. Performance of ChatGPT on a radiology board-style examination: Insights into current strengths and limitations. Radiology, 307(5):e230582, 2023. doi: 10.1148/radiol.230582.

[10] Diogo Almeida. Introducing System One models & Jev. https://typesafe.ai/blog/introducing-system-one-models-and-jev, 2026. TypeSafe blog, 15 September 2026.

[11] TypeSafe. System One API documentation. https://docs.typesafe.ai/api, 2026. Accessed 1 October 2026.

[12] Archer Hume. Jev’s architecture unmasked. https://archerhume.com/posts/jevs-architecture-unmasked, 2026. 17 September 2026.

[13] OpenAI. Decisions. https://developers.openai.com/api/docs/guides/decisions, 2026. API guide; public beta with gpt-6-luna released 6 October 2026; accessed 6 October 2026.

[14] Qwen Team. Qwen3.8-27B. https://huggingface.co/Qwen/Qwen3.8-27B, 2026. Apache-2.0; revision 1d4bf0f.

[15] Jared Palmer. Kev: a family of decision models built on Qwen3.5 and Qwen3.8. https://github.com/jaredpalmer/kev, 2026. Apache-2.0.

[16] Alfredo Madrid-García and Beatriz Merino-Barbancho. Jev in medicine: A benchmark evaluation, 2026. arXiv:2609.34024.

[17] Jiaju Huang, Hao Yang, Xinyu Ma, Xinglong Liang, Kunyan Cai, Junqiang Ma, Shaobin Chen, Yue Sun, and Tao Tan. Can Jev judge radiology reports? Evaluating a System One model for clinical factuality, 2026. arXiv:2609.27607.

[18] Suchin Gururangan, Ana Marasović, Swabha Swayamdipta, Kyle Lo, Iz Beltagy, Doug Downey, and Noah A. Smith. Don’t stop pretraining: Adapt language models to domains and tasks. In Proceedings of the 58th Annual Meeting of the Association for Computational Linguistics, 2020. arXiv:2004.10964.

[19] Harsha Nori, Yin Tat Lee, Sheng Zhang, Dean Carignan, Richard Edgar, Nicolo Fusi, Nicholas King, Jonathan Larson, Yuanzhi Li, Weishung Liu, Renqian Luo, Scott Mayer McKinney, Robert Osazuwa Ness, Hoifung Poon, Tao Qin, Naoto Usuyama, Chris White, and Eric Horvitz. Can generalist foundation models outcompete special-purpose tuning? Case study in medicine, 2023. arXiv:2311.16452.

[20] Zexiao Wang, Zihao Zhang, Xudong Wang, Pan Wang, Ziyi Ye, Haoyu Zhao, Zuxuan Wu, and Shuicheng Yan. Chinese-Jev: Bringing System One model to Chinese-language tasks, 2026. arXiv:2609.36965.

[21] Jean-Benoit Delbrouck, Pierre Chambon, Zhihong Chen, Maya Varma, Andrew Johnston, Louis Blankemeier, Dave Van Veen, Tan Bui, Steven Truong, and Curtis Langlotz. RadGraph-XL: A large-scale expert-annotated dataset for entity and relation extraction from radiology reports. In Findings of the Association for Computational Linguistics: ACL 2024, pages 12902–12915. Association for Computational Linguistics, 2024. doi: 10.18653/v1/2024.findings-acl.765.

[22] Dina Demner-Fushman et al. Preparing a collection of radiology examinations for distribution and retrieval. Journal of the American Medical Informatics Association, 23(2):304–310, 2016. doi: 10.1093/jamia/ocv080.

[23] Ibrahim Ethem Hamamci et al. Generalist foundation models from a multimodal dataset for 3D computed tomography. Nature Biomedical Engineering, 10(8):1610–1628, 2026. doi: 10.1038/s41551-025-01599-y.

[24] Vishwanatha M. Rao, Serena Zhang, Julián N. Acosta, Subathra Adithan, and Pranav Rajpurkar. ReX-Err: Synthesizing clinically meaningful errors in diagnostic radiology reports. In Biocomputing 2025: Proceedings of the Pacific Symposium, pages 70–81. World Scientific, 2025. doi: 10.1142/9789819807024_0006.

[25] Alistair E. W. Johnson, Tom J. Pollard, Seth J. Berkowitz, Nathaniel R. Greenbaum, Matthew P. Lungren, Chih-ying Deng, Roger G. Mark, and Steven Horng. MIMIC-CXR, a de-identified publicly available database of chest radiographs with free-text reports. Scientific Data, 6:317, 2019. doi: 10.1038/s41597-019-0322-0.

[26] European Society of Radiology. Eurorad: radiological case database. https://www.eurorad.org, n.d. CC BY-NC-SA 4.0.

[27] WangLab. eurorad-reasoning. https://huggingface.co/datasets/wanglab/eurorad-reasoning, 2025. Hugging Face dataset; 3,669 Eurorad cases.

[28] Sebastian Wind, Jeta Sopa, Daniel Truhn, Mahshad Lotfinia, Tri-Thien Nguyen, Keno Bressem, Lisa Adams, Mirabela Rusu, Harald Köstler, Gerhard Wellein, Andreas Maier, and Soroosh Tayebi Arasteh. Multi-step retrieval and reasoning improves radiology question answering with large language models. npj Digital Medicine, 8:790, 2025. doi: 10.1038/s41746-025-02250-5.

[29] Hanjie Chen, Zhouxiang Fang, Yash Singla, and Mark Dredze. Benchmarking large language models on answering and explaining challenging medical questions. arXiv preprint arXiv:2402.18060, 2024.

[30] Ankit Pal, Logesh Kumar Umapathi, and Malaikannan Sankarasubbu. MedMCQA: A large-scale multi-subject multi-choice dataset for medical domain question answering. In Proceedings of the Conference on Health, Inference, and Learning, volume 174 of PMLR, pages 248–260, 2022. arXiv:2203.14371.

[31] Di Jin, Eileen Pan, Nassim Oufattole, Wei-Hung Weng, Hanyi Fang, and Peter Szolovits. What disease does this patient have? A large-scale open domain question answering dataset from medical exams. Applied Sciences, 11(14):6421, 2021. doi: 10.3390/app11146421.

[32] Dan Hendrycks, Collin Burns, Steven Basart, Andy Zou, Mantas Mazeika, Dawn Song, and Jacob Steinhardt. Measuring massive multitask language understanding. In International Conference on Learning Representations, 2021. arXiv:2009.03300.

[33] Qiao Jin, Bhuwan Dhingra, Zhengping Liu, William W. Cohen, and Xinghua Lu. PubMedQA: A dataset for biomedical research question answering. In Proceedings of the 2019 Conference on Empirical Methods in Natural Language Processing, pages 2567–2577, 2019. doi: 10.18653/v1/D19-1259.

[34] Yuxin Zuo, Shang Qu, Yifei Li, Zhang-Ren Chen, Xuekai Zhu, Ermo Hua, Kaiyan Zhang, Ning Ding, and Bowen Zhou. MedXpertQA: Benchmarking expert-level medical reasoning and understanding. In Proceedings of the 42nd International Conference on Machine Learning, volume 267 of PMLR, pages 80961–80990, 2025. arXiv:2501.18362.

[35] Geoffrey Hinton, Oriol Vinyals, and Jeff Dean. Distilling the knowledge in a neural network, 2015. arXiv:1503.02531.

[36] Pierre Chambon, Jean-Benoit Delbrouck, Thomas Sounack, Shih-Cheng Huang, Zhihong Chen, Maya Varma, Steven QH Truong, Chu The Chuong, and Curtis P. Langlotz. CheXpert Plus: Augmenting a large chest x-ray dataset with text radiology reports, patient demographics and additional image formats, 2024. arXiv:2405.19538.

[37] Xiaoman Zhang, Julián N. Acosta, Josh Miller, Ouwen Huang, and Pranav Rajpurkar. ReXGradient-160K: A large-scale publicly available dataset of chest radiographs with free-text reports, 2025. arXiv:2505.00228.

[38] Andrew Sellergren, Sahar Kazemzadeh, Tiam Jaroensri, Atilla Kiraly, Madeleine Traverse, et al. MedGemma technical report, 2025. arXiv:2507.05201.

[39] Jeremy Irvin, Pranav Rajpurkar, Michael Ko, Yifan Yu, Silviana Ciurea-Ilcus, Chris Chute, Henrik Marklund, Behzad Haghgoo, Robyn Ball, Katie Shpanskaya, et al. CheXpert: A large chest radiograph dataset with uncertainty labels and expert comparison. In Proceedings of the AAAI Conference on Artificial Intelligence, 2019. arXiv:1901.07031.

[40] Fabrizio Gilardi, Meysam Alizadeh, and Maël Kubli. ChatGPT outperforms crowd workers for text-annotation tasks. Proceedings of the National Academy of Sciences, 120(30):e2305016120, 2023. doi: 10.1073/pnas.2305016120.

[41] Carla E. Brodley and Mark A. Friedl. Identifying mislabeled training data. Journal of Artificial Intelligence Research, 11:131–167, 1999. doi: 10.1613/jair.606.

[42] Nicholas Pangakis, Samuel Wolken, and Neil Fasching. Automated annotation with generative AI requires validation, 2023. arXiv:2306.00176.

[43] Edward J. Hu et al. LoRA: Low-rank adaptation of large language models. In International Conference on Learning Representations, 2022. arXiv:2106.09685.

[44] Oriol Vinyals, Meire Fortunato, and Navdeep Jaitly. Pointer networks. In Advances in Neural Information Processing Systems, volume 28, 2015. arXiv:1506.03134.

[45] Chuan Guo, Geoff Pleiss, Yu Sun, and Kilian Q. Weinberger. On calibration of modern neural networks. In Proceedings of the 34th International Conference on Machine Learning, volume 70 of PMLR, pages 1321–1330, 2017. arXiv:1706.04599.

[46] Marco Saerens, Patrice Latinne, and Christine Decaestecker. Adjusting the outputs of a classifier to new a priori probabilities: A simple procedure. Neural Computation, 14(1):21–41, 2002. doi: 10.1162/089976602753284446.

[47] Bradley Efron and Robert J. Tibshirani. An Introduction to the Bootstrap. Chapman & Hall, 1993.

[48] Sture Holm. A simple sequentially rejective multiple test procedure. Scandinavian Journal of Statistics, 6(2):65–70, 1979.

[49] Dave Van Veen, Cara Van Uden, Louis Blankemeier, Jean-Benoit Delbrouck, Asad Aali, Christian Bluethgen, Anuj Pareek, Malgorzata Polacin, Eduardo Pontes Reis, Anna Seehofnerová, Nidhi Rohatgi, Poonam Hosamani, William Collins, Neera Ahuja, Curtis P. Langlotz, Jason Hom, Sergios Gatidis, John Pauly, and Akshay S. Chaudhari. Adapted large language models can outperform medical experts in clinical text summarization. Nature Medicine, 30(4):1134–1142, 2024. doi: 10.1038/s41591-024-02855-5.

[50] Merel Huisman and Nika Rasoolzadeh. LLMs for radiology reports: From general purpose to light-weight domain adaptation. Radiology, 316(2):e252524, 2025. doi: 10.1148/radiol.252524.

[51] Christian Bluethgen, Dave Van Veen, Cyril Zakka, Katherine E. Link, Aaron Hunter Fanous, Roxana Daneshjou, Thomas Frauenfelder, Curtis P. Langlotz, Sergios Gatidis, and Akshay Chaudhari. Best practices for large language models in radiology. Radiology, 315(1):e240528, 2025. doi: 10.1148/radiol.240528.

[52] Cody H. Savage, Adway Kanhere, Vishwa Parekh, Curtis P. Langlotz, Anupam Joshi, Heng Huang, and Florence X. Doo. Open-source large language models in radiology: A review and tutorial for practical research and clinical deployment. Radiology, 314(1):e241073, 2025. doi: 10.1148/radiol.241073.

[53] Magdalini Paschali, Zhihong Chen, Louis Blankemeier, Maya Varma, Alaa Youssef, Christian Bluethgen, Curtis Langlotz, Sergios Gatidis, and Akshay Chaudhari. Foundation models in radiology: What, how, why, and why not. Radiology, 314(2):e240597, 2025. doi: 10.1148/radiol.240597.

[54] Yu Gu, Robert Tinn, Hao Cheng, Michael Lucas, Naoto Usuyama, Xiaodong Liu, Tristan Naumann, Jianfeng Gao, and Hoifung Poon. Domain-specific language model pretraining for biomedical natural language processing. ACM Transactions on Computing for Healthcare, 3(1):1–23, 2021. doi: 10.1145/3458754.

