## Supplementary for "RadKev: An Open-Weight Decision Model for Radiology"

**Table S1.** Teacher-labeled question kinds. Candidates were answered by MedGemma-27B-text and Qwen3.8-27B; a question was retained when both ranked the same answer first. Order questions saw only the clinical indication. First run, labels used for training, model selection and the main evaluation; regenerated, labels after rescoring MedGemma with the pre-filled reasoning segment.

| | | | | | Retained (%) | |
|---|---|---|---|---|---|---|
| Question kind | Input | Answer type | Answers | Candidates | First run | Regenerated |
| Critical finding | Report | Yes/no | 2 | 3,078 | 2,201 (71.5) | 2,373 (77.1) |
| Urgency | Report | Ordinal | 4 | 2,985 | 2,365 (79.2) | 2,339 (78.4) |
| Follow-up | Report | Choice | 5 | 3,097 | 1,923 (62.1) | 1,920 (62.0) |
| Interval change | Report with a comparison | Choice | 5 | 1,224 | 983 (80.3) | 998 (81.5) |
| Incidental finding | Report | Yes/no | 2 | 3,116 | 2,114 (67.8) | 2,258 (72.5) |
| Finding status | Chest radiograph report | Choice | 4 | 3,000 | 2,540 (84.7) | 2,615 (87.2) |
| First examination | Clinical indication | Choice | 14 | 3,968 | 3,058 (77.1) | 3,152 (79.4) |
| Contrast | Clinical indication | Choice | 4 | 3,989 | 1,121 (28.1) | 909 (22.8) |
| Priority | Clinical indication | Ordinal | 3 | 4,043 | 2,644 (65.4) | 2,656 (65.7) |
| Appropriateness | Indication and ordered examination | Ordinal | 3 | 4,500 | 3,608 (80.2) | 3,387 (75.3) |
| Total | | | | 33,000 | 22,557 (68.4) | 22,607 (68.5) |

### Supplementary Note S3. Training details

**Augmentation.** Each multiple-choice question with a single correct answer received at most one modification: the correct option was replaced by a "none of the above" option, which then became the correct answer (probability 0.10; questions with more than two options); a "none of the above" option was added as an incorrect alternative (probability 0.12); or an irrelevant distractor was added (probability 0.15). With probability 0.25, a record additionally contributed a minimal pair in which the same question was posed with a "none of the above" option, once with and once without its correct option. With the 1,000 replayed Kev records, training comprised 36,954 records and 44,068 examples for RadKev-27B (44,070 for RadKev-9B), including the minimal pairs.

### Supplementary Tables

**Table S2.** Primary comparison by task: accuracy (%) of Kev-27B and RadKev-27B and their difference in percentage points (95% confidence interval, paired cluster bootstrap with 2,000 resamples). Holm $p$: adjusted across the fifteen benchmark tasks. Top: tasks with human-labeled ground truth; middle: ReXErr, with ground truth known by construction; bottom: averages (the first row is the prespecified primary outcome).

| Task | n | Kev-27B | RadKev-27B | Δ (95% CI) | Holm $p$ |
|---|---|---|---|---|---|
| IU: finding present | 5,696 | 96.6 | 96.8 | 0.2 (−0.1 to 0.5) | 1.000 |
| IU: normal study | 2,848 | 87.6 | 90.2 | 2.6 (1.6 to 3.5) | <0.001 |
| IU: which finding | 674 | 95.4 | 96.9 | 1.5 (0.3 to 2.7) | 0.120 |
| Eurorad: diagnosis | 346 | 82.7 | 94.2 | 11.6 (8.1 to 15.6) | <0.001 |
| Eurorad: subspecialty classification | 185 | 84.3 | 85.4 | 1.1 (−2.2 to 4.5) | 1.000 |
| RSNA-RadioQA: diagnosis | 79 | 73.4 | 75.9 | 2.5 (−2.5 to 7.6) | 1.000 |
| RadCases: ACR panel | 132 | 66.7 | 67.4 | 0.8 (−7.6 to 9.6) | 1.000 |
| RadCases: ACR topic | 95 | 61.1 | 63.2 | 2.1 (−2.0 to 6.6) | 1.000 |
| MedMCQA: radiology | 69 | 72.5 | 78.3 | 5.8 (−4.6 to 16.7) | 1.000 |
| MedMCQA: other subjects | 151 | 64.9 | 76.2 | 11.3 (5.7 to 17.3) | <0.001 |
| MedQA | 331 | 84.0 | 87.9 | 3.9 (0.9 to 6.9) | 0.120 |
| MedXpertQA | 641 | 26.4 | 30.6 | 4.2 (1.2 to 7.3) | 0.066 |
| MMLU | 79 | 92.4 | 94.9 | 2.5 (0.0 to 6.3) | 1.000 |
| PubMedQA | 108 | 78.7 | 79.6 | 0.9 (−1.9 to 4.6) | 1.000 |
| ReXErr: report error | 2,708 | 72.1 | 94.7 | 22.6 (20.9 to 24.3) | <0.001 |
| Task mean, all tasks (primary) | 14,142 | 75.9 | 80.8 | 4.9 (3.8 to 6.2) | – |
| Task mean, human-labeled | 11,434 | 76.2 | 79.8 | 3.6 (2.4 to 5.0) | – |
| All questions pooled | 14,142 | 84.8 | 90.6 | 5.8 (5.3 to 6.3) | – |
| Human-labeled questions pooled | 11,434 | 87.8 | 89.6 | 1.8 (1.4 to 2.2) | – |

**Table S3.** Accuracy (%) of every system on every benchmark task and, at the bottom, agreement with the CT-RATE classifier labels, which are not part of the benchmark. Qwen, Qwen3.8-27B, which was not scored on the RadCases topic question (225 options); a dash marks a value not computed.

| Task | RadKev-27B | RadKev-9B | Kev-27B | Kev-9B | Qwen | OpenAI | Jev |
|---|---|---|---|---|---|---|---|
| IU: finding present | 96.8 | 98.0 | 96.6 | 97.1 | 96.6 | 96.3 | 97.0 |
| IU: normal study | 90.2 | 92.6 | 87.6 | 91.7 | 93.6 | 89.8 | 87.9 |
| IU: which finding | 96.9 | 96.6 | 95.4 | 95.8 | 96.1 | 95.5 | 96.4 |
| Eurorad: diagnosis | 94.2 | 90.5 | 82.7 | 75.1 | 84.4 | 84.7 | 81.2 |
| Eurorad: subspecialty classification | 85.4 | 84.3 | 84.3 | 77.8 | 87.0 | 78.9 | 82.2 |
| RSNA-RadioQA: diagnosis | 75.9 | 69.6 | 73.4 | 60.8 | 77.2 | 75.9 | 73.4 |
| RadCases: ACR panel | 67.4 | 68.2 | 66.7 | 75.0 | 55.3 | 64.4 | 62.9 |
| RadCases: ACR topic | 63.2 | 61.1 | 61.1 | 58.9 | – | 29.5 | 63.2 |
| MedMCQA: radiology | 78.3 | 69.6 | 72.5 | 59.4 | 75.4 | 78.3 | 76.8 |
| MedMCQA: other subjects | 76.2 | 65.6 | 64.9 | 53.6 | 67.5 | 76.8 | 75.5 |
| MedQA | 87.9 | 73.1 | 84.0 | 70.1 | 84.6 | 88.2 | 87.0 |
| MedXpertQA | 30.6 | 21.7 | 26.4 | 16.7 | 31.4 | 40.6 | 31.7 |
| MMLU | 94.9 | 91.1 | 92.4 | 87.3 | 92.4 | 93.7 | 97.5 |
| PubMedQA | 79.6 | 79.6 | 78.7 | 75.0 | 73.1 | 68.5 | 78.7 |
| ReXErr: report error | 94.7 | 92.9 | 72.1 | 56.9 | 69.1 | 78.5 | 75.6 |
| Task mean, all tasks | 80.8 | 77.0 | 75.9 | 70.1 | – | 76.0 | 77.8 |
| Task mean, human-labeled | 79.8 | 75.8 | 76.2 | 71.0 | – | 75.8 | 78.0 |
| CT-RATE: finding present | 98.7 | 98.3 | 94.9 | 91.5 | 95.6 | – | – |
| CT-RATE: normal study | 98.7 | 98.8 | 96.3 | 96.6 | 96.4 | – | – |
| CT-RATE: which finding | 99.0 | 99.1 | 96.1 | 97.1 | 96.4 | – | – |

**Table S4.** Paired comparisons in percentage points (95% confidence interval): task-averaged accuracy over all benchmark tasks and over the tasks with human-labeled ground truth, and accuracy pooled over all questions. Comparisons with an LLM use the questions with at most 16 options. Tasks: number of tasks compared.

| Comparison | Task mean, all | Task mean, human | Pooled, all | Tasks |
|---|---|---|---|---|
| RadKev-27B − Kev-27B | 4.9 (3.8 to 6.2) | 3.6 (2.4 to 5.0) | 5.8 (5.3 to 6.3) | 15 |
| RadKev-9B − Kev-9B | 6.9 (5.5 to 8.4) | 4.8 (3.3 to 6.4) | 8.5 (8.0 to 9.0) | 15 |
| RadKev-27B − Qwen3.8-27B | 4.7 (3.4 to 6.0) | 3.0 (1.6 to 4.5) | 4.9 (4.4 to 5.4) | 14 |
| RadKev-9B − Qwen3.8-27B | 0.7 (−1.0 to 2.5) | −1.1 (−3.0 to 0.8) | 4.4 (3.9 to 5.0) | 14 |
| RadKev-27B − RadKev-9B | 3.9 (2.4 to 5.4) | 4.0 (2.5 to 5.6) | 0.4 (0.1 to 0.8) | 15 |
| Kev-27B − Kev-9B | 5.8 (4.2 to 7.5) | 5.1 (3.4 to 6.9) | 3.1 (2.7 to 3.6) | 15 |
| RadKev-9B − Kev-27B | 1.1 (−0.5 to 2.7) | −0.4 (−2.1 to 1.5) | 5.3 (4.8 to 5.9) | 15 |
| Qwen3.8-27B − Kev-27B | 0.4 (−0.8 to 1.7) | 0.7 (−0.6 to 2.1) | 0.9 (0.3 to 1.5) | 14 |
| RadKev-27B − OpenAI Decisions | 4.8 (3.2 to 6.4) | 4.0 (2.3 to 5.7) | 3.6 (3.1 to 4.1) | 15 |
| RadKev-9B − OpenAI Decisions | 1.0 (−0.8 to 2.9) | 0.0 (−1.9 to 2.0) | 3.2 (2.7 to 3.7) | 15 |
| OpenAI Decisions − Kev-27B | 0.1 (−1.5 to 1.7) | −0.4 (−2.0 to 1.4) | 2.1 (1.6 to 2.7) | 15 |
| OpenAI Decisions − Qwen3.8-27B | 1.9 (0.5 to 3.4) | 1.3 (−0.2 to 2.9) | 1.4 (1.0 to 1.9) | 14 |
| RadKev-27B − Jev | 3.0 (1.7 to 4.4) | 1.9 (0.4 to 3.3) | 4.4 (4.0 to 5.0) | 15 |
| RadKev-9B − Jev | −0.8 (−2.5 to 0.9) | −2.1 (−3.8 to −0.3) | 4.0 (3.5 to 4.6) | 15 |
| Jev − Kev-27B | 1.9 (0.3 to 3.4) | 1.8 (0.2 to 3.4) | 1.3 (0.8 to 1.9) | 15 |
| Jev − Qwen3.8-27B | 1.4 (0.0 to 2.8) | 1.0 (−0.5 to 2.5) | 0.4 (−0.1 to 0.9) | 14 |
| OpenAI Decisions − Jev | −1.8 (−3.4 to −0.3) | −2.2 (−3.9 to −0.5) | 0.8 (0.4 to 1.2) | 15 |

**Table S5.** Accuracy (%, 95% confidence interval) on the reasoning sample of 1,675 benchmark questions, pooled over questions and averaged over tasks, with the questions of the six examination tasks pooled into one task, and with reasoning disabled and enabled for Qwen3.8-27B.

| System | Pooled over questions | Averaged over tasks |
|---|---|---|
| RadKev-27B | 89.0 (87.6 to 90.4) | 86.4 (84.6 to 88.2) |
| Kev-27B | 82.0 (80.2 to 83.8) | 80.4 (78.4 to 82.5) |
| RadKev-9B | 87.1 (85.6 to 88.6) | 84.2 (82.3 to 86.2) |
| Qwen3.8-27B | 83.6 (81.9 to 85.2) | 81.8 (79.8 to 83.8) |
| Qwen3.8-27B, reasoning | 85.1 (83.5 to 86.8) | 83.5 (81.6 to 85.4) |

**Table S6.** Latency per question on 60 benchmark records (one request at a time, two RTX A6000 GPUs, Hugging Face Transformers, bfloat16): median and 95th percentile. $n$: timed records for the decision models, including the OpenAI Decisions API and Jev, whose latency per question is the latency of the record divided by its number of questions, and timed questions for Qwen3.8-27B.

| System | Mode | Median (ms) | 95th percentile (ms) | $n$ |
|---|---|---|---|---|
| RadKev-27B | one pass per record | 128 | 322 | 55 |
| Kev-27B | one pass per record | 129 | 278 | 55 |
| RadKev-9B | one pass per record | 43 | 112 | 55 |
| Kev-9B | one pass per record | 43 | 86 | 55 |
| OpenAI Decisions | end to end, 4 in flight | 145 | 212 | 7,471 |
| OpenAI Decisions | end to end, one at a time | 175 | 276 | 27 |
| Jev | end to end, 4 in flight | 168 | 253 | 7,471 |
| Jev | end to end, one at a time | 185 | 259 | 27 |
| Qwen3.8-27B | option-letter logits | 145 | 245 | 99 |
| Qwen3.8-27B | generated letter | 234 | 332 | 99 |
| Qwen3.8-27B | reasoning | 16906 | 179802 | 54 |

**Table S7.** Kev's out-of-domain transfer suite (656 questions): accuracy (%) of each specialized model and of the released model it started from, and their difference in percentage points (95% confidence interval).

| Comparison | RadKev | Kev | Δ (95% CI) |
|---|---|---|---|
| RadKev-9B vs Kev-9B | 83.7 | 82.2 | 1.5 (0.0 to 3.1) |
| RadKev-27B vs Kev-27B | 84.5 | 85.1 | −0.6 (−2.3 to 1.2) |

**Table S8.** Calibration and selective prediction on the human-labeled questions: expected calibration error (ECE) over ten equal-width bins, Brier score, confident errors (% of questions answered incorrectly with a confidence of at least 0.9) and coverage at a 5% error budget (%), as scored and after two-fold cross-fitted temperature recalibration on the test split (recal.). The OpenAI Decisions API and Jev return probabilities rounded to two decimals, so they are reported as scored only.

| System | ECE | ECE (recal.) | Brier | Conf. err. | Conf. err. (recal.) | Cov. 5% | Cov. 5% (recal.) |
|---|---|---|---|---|---|---|---|
| RadKev-27B | 0.015 | 0.015 | 0.149 | 1.4 | 0.8 | 88.4 | 88.2 |
| RadKev-9B | 0.021 | 0.024 | 0.148 | 1.1 | 0.8 | 89.4 | 89.7 |
| Kev-27B | 0.018 | 0.015 | 0.164 | 0.6 | 0.8 | 85.5 | 85.3 |
| Kev-9B | 0.023 | 0.017 | 0.170 | 0.7 | 1.1 | 86.3 | 86.1 |
| Qwen3.8-27B | 0.022 | 0.022 | 0.154 | 2.0 | 0.9 | 86.6 | 88.4 |
| OpenAI Decisions | 0.019 | – | 0.163 | 2.4 | – | 85.9 | – |
| Jev | 0.031 | – | 0.161 | 1.4 | – | 86.8 | – |

**Table S9.** Eurorad subspecialty classification with the full state and with the imaging findings removed (age, sex and clinical history retained), as before the read: accuracy (%) and change in percentage points (95% confidence interval), 185 questions. Accuracies with the full state were recomputed in this analysis, with each question in its own request; they differ from the main evaluation by one question for Kev-27B and two for Qwen3.8-27B.

| System | Full state | Findings removed | Change (95% CI) |
|---|---|---|---|
| RadKev-27B | 85.4 | 73.0 | −12.4 (−18.4 to −7.0) |
| Kev-27B | 83.8 | 70.3 | −13.5 (−19.5 to −8.1) |
| RadKev-9B | 84.3 | 70.3 | −14.1 (−20.0 to −8.1) |
| Kev-9B | 77.8 | 65.9 | −11.9 (−18.4 to −4.9) |
| Qwen3.8-27B | 88.1 | 70.3 | −17.8 (−23.8 to −11.9) |

**Table S10.** Options-only control: accuracy (%, 95% confidence interval) with the full case and with only the question and its options shown. Accuracies with the full case were recomputed in this analysis; for Qwen3.8-27B they differ from the main evaluation by less than 0.5 percentage points because of batch padding.

| Task | System | Full case | Options only |
|---|---|---|---|
| Eurorad diagnosis | RadKev-27B | 94.2 (91.6 to 96.5) | 76.9 (72.3 to 81.2) |
| | Kev-27B | 82.7 (78.3 to 86.4) | 47.7 (42.5 to 52.9) |
| | RadKev-9B | 90.5 (87.0 to 93.4) | 81.5 (77.5 to 85.5) |
| | Kev-9B | 75.1 (70.5 to 79.5) | 38.4 (33.2 to 43.9) |
| | Qwen3.8-27B | 84.1 (80.1 to 87.9) | 50.0 (45.1 to 55.5) |
| RSNA-RadioQA | RadKev-27B | 75.9 (67.1 to 84.8) | 50.6 (39.2 to 62.0) |
| | Kev-27B | 73.4 (63.3 to 82.3) | 40.5 (30.3 to 50.6) |
| | RadKev-9B | 69.6 (59.5 to 79.7) | 48.1 (38.0 to 59.5) |
| | Kev-9B | 60.8 (49.4 to 70.9) | 41.8 (30.4 to 53.2) |
| | Qwen3.8-27B | 77.2 (68.4 to 86.1) | 46.8 (36.7 to 58.2) |

**Table S11.** Accuracy (%) under each answer space on 30 Eurorad diagnosis and 30 RSNA-RadioQA questions; K, number of options offered. Random and most similar: alternatives drawn from the pool of diagnoses at random or in order of similarity to the correct answer. LLM-written: alternatives written by Qwen3.8-27B; with 16 options, 18 Eurorad and 21 RSNA-RadioQA questions retained enough alternatives after filtering. Original + LLM-written: the original options supplemented with up to 8 LLM-written alternatives. Qwen3.8-27B generated the number of the chosen option.

| Source | Alternatives | K | RadKev-27B | Kev-27B | RadKev-9B | Kev-9B | Qwen | OpenAI | Jev |
|---|---|---|---|---|---|---|---|---|---|
| Eurorad diagnosis | Original options | – | 96.7 | 90.0 | 100.0 | 80.0 | 90.0 | 96.7 | 96.7 |
| | Random | 2 | 100.0 | 96.7 | 100.0 | 96.7 | 96.7 | 100.0 | 100.0 |
| | | 4 | 100.0 | 96.7 | 100.0 | 96.7 | 96.7 | 100.0 | 100.0 |
| | | 16 | 100.0 | 100.0 | 96.7 | 93.3 | 96.7 | 96.7 | 100.0 |
| | | 64 | 93.3 | 100.0 | 96.7 | 93.3 | 100.0 | 90.0 | 96.7 |
| | | 255 | 93.3 | 93.3 | 93.3 | 90.0 | 93.3 | 60.0 | 96.7 |
| | Most similar | 2 | 93.3 | 96.7 | 90.0 | 90.0 | 96.7 | 93.3 | 96.7 |
| | | 4 | 93.3 | 93.3 | 80.0 | 76.7 | 90.0 | 93.3 | 90.0 |
| | | 16 | 93.3 | 90.0 | 80.0 | 73.3 | 83.3 | 90.0 | 83.3 |
| | | 64 | 80.0 | 86.7 | 70.0 | 70.0 | 76.7 | 66.7 | 83.3 |
| | | 255 | 73.3 | 80.0 | 63.3 | 73.3 | 80.0 | 46.7 | 80.0 |
| | LLM-written | 2 | 100.0 | 100.0 | 96.7 | 86.7 | 100.0 | 100.0 | 100.0 |
| | | 4 | 96.7 | 96.7 | 96.7 | 90.0 | 96.7 | 96.7 | 100.0 |
| | | 8 | 93.3 | 93.3 | 96.7 | 86.7 | 93.3 | 93.3 | 96.7 |
| | | 16 | 100.0 | 94.4 | 94.4 | 72.2 | 100.0 | 94.4 | 94.4 |
| | Original + LLM-written | – | 90.0 | 83.3 | 90.0 | 80.0 | 83.3 | 83.3 | 93.3 |
| RSNA-RadioQA | Original options | – | 73.3 | 73.3 | 63.3 | 60.0 | 70.0 | 76.7 | 70.0 |
| | Random | 2 | 100.0 | 100.0 | 96.7 | 96.7 | 100.0 | 100.0 | 100.0 |
| | | 4 | 96.7 | 96.7 | 90.0 | 93.3 | 100.0 | 96.7 | 96.7 |
| | | 16 | 90.0 | 90.0 | 90.0 | 86.7 | 93.3 | 83.3 | 86.7 |
| | | 64 | 86.7 | 90.0 | 90.0 | 90.0 | 90.0 | 80.0 | 86.7 |
| | | 255 | 83.3 | 76.7 | 83.3 | 80.0 | 80.0 | 60.0 | 73.3 |
| | Most similar | 2 | 73.3 | 76.7 | 63.3 | 63.3 | 86.7 | 76.7 | 70.0 |
| | | 4 | 63.3 | 66.7 | 60.0 | 63.3 | 80.0 | 70.0 | 66.7 |
| | | 16 | 60.0 | 63.3 | 36.7 | 43.3 | 43.3 | 56.7 | 56.7 |
| | | 64 | 53.3 | 53.3 | 40.0 | 36.7 | 56.7 | 43.3 | 53.3 |
| | | 255 | 33.3 | 43.3 | 26.7 | 26.7 | 36.7 | 23.3 | 46.7 |
| | LLM-written | 2 | 86.7 | 86.7 | 86.7 | 80.0 | 86.7 | 83.3 | 86.7 |
| | | 4 | 83.3 | 80.0 | 80.0 | 66.7 | 80.0 | 80.0 | 76.7 |
| | | 8 | 80.0 | 73.3 | 76.7 | 63.3 | 76.7 | 73.3 | 70.0 |
| | | 16 | 81.0 | 76.2 | 61.9 | 57.1 | 71.4 | 76.2 | 66.7 |
| | Original + LLM-written | – | 76.7 | 70.0 | 63.3 | 56.7 | 66.7 | 73.3 | 66.7 |

**Table S12.** External test set (RadGraph-XL): accuracy (%, 95% confidence interval) and paired differences in percentage points on all 4,037 status questions, on the 2,991 questions about definite findings (present or absent; uncertain option removed from every answer) and on the 1,046 hedged findings (annotated as uncertain).

| System or comparison | All questions | Definite findings | Hedged findings |
|---|---|---|---|
| RadKev-27B | 76.3 (75.2 to 77.4) | 90.2 (89.2 to 91.3) | 39.1 (36.1 to 42.1) |
| RadKev-9B | 72.5 (71.3 to 73.6) | 87.5 (86.3 to 88.7) | 33.2 (30.3 to 36.1) |
| Kev-27B | 77.4 (76.2 to 78.5) | 91.3 (90.3 to 92.3) | 40.7 (37.7 to 43.8) |
| Kev-9B | 70.7 (69.5 to 71.9) | 85.2 (83.9 to 86.4) | 35.9 (33.0 to 38.7) |
| Qwen3.8-27B | 78.4 (77.2 to 79.6) | 90.3 (89.2 to 91.4) | 55.4 (52.4 to 58.4) |
| OpenAI Decisions | 69.7 (68.4 to 70.9) | 81.9 (80.6 to 83.4) | 36.8 (33.7 to 39.8) |
| Jev | 76.6 (75.5 to 77.7) | 89.3 (88.2 to 90.4) | 43.1 (40.0 to 46.1) |
| RadKev-27B − Kev-27B | −1.1 (−1.6 to −0.6) | −1.1 (−1.6 to −0.6) | −1.6 (−3.2 to −0.1) |
| RadKev-9B − Kev-9B | 1.8 (1.0 to 2.6) | 2.4 (1.5 to 3.3) | −2.7 (−4.2 to −1.1) |
| RadKev-27B − Qwen3.8-27B | −2.2 (−3.0 to −1.3) | −0.1 (−0.8 to 0.7) | −16.3 (−18.6 to −14.0) |
| RadKev-27B − Jev | −0.3 (−1.0 to 0.5) | 0.9 (0.3 to 1.6) | −4.0 (−6.3 to −1.8) |
| RadKev-27B − OpenAI Decisions | 6.6 (5.7 to 7.6) | 8.3 (7.3 to 9.3) | 2.3 (0.1 to 4.5) |
| RadKev-9B − Qwen3.8-27B | −6.0 (−7.0 to −5.0) | −2.8 (−3.7 to −1.9) | −22.2 (−24.9 to −19.5) |
